# Powerful mapping of *cis*-genetic effects on gene expression across diverse populations reveals novel disease-critical genes

**DOI:** 10.1101/2024.09.25.24314410

**Authors:** Kai Akamatsu, Stephen Golzari, Tiffany Amariuta

## Abstract

While disease-associated variants identified by genome-wide association studies (GWAS) most likely regulate gene expression levels, linking variants to target genes is critical to determining the functional mechanisms of these variants. Genetic effects on gene expression have been extensively characterized by expression quantitative trait loci (eQTL) studies, yet data from non-European populations is limited. This restricts our understanding of disease to genes whose regulatory variants are common in European populations. While previous work has leveraged data from multiple populations to improve GWAS power and polygenic risk score (PRS) accuracy, multi-ancestry data has not yet been used to better estimate *cis*-genetic effects on gene expression. Here, we present a new method, Multi-Ancestry Gene Expression Prediction Regularized Optimization (MAGEPRO), which constructs robust genetic models of gene expression in understudied populations or cell types by fitting a regularized linear combination of eQTL summary data across diverse cohorts. In simulations, our tool generates more accurate models of gene expression than widely-used LASSO and the state-of-the-art multi-ancestry PRS method, PRS-CSx, adapted to gene expression prediction. We attribute this improvement to MAGEPRO’s ability to more accurately estimate causal eQTL effect sizes (*p* < 3.98 × 10^-4^, two-sided paired t-test). With real data, we applied MAGEPRO to 8 eQTL cohorts representing 3 ancestries (average *n* = 355) and consistently outperformed each of 6 competing methods in gene expression prediction tasks. Integration with GWAS summary statistics across 66 complex traits (representing 22 phenotypes and 3 ancestries) resulted in 2,331 new gene-trait associations, many of which replicate across multiple ancestries, including *PHTF1* linked to white blood cell count, a gene which is overexpressed in leukemia patients. MAGEPRO also identified biologically plausible novel findings, such as *PIGB*, an essential component of GPI biosynthesis, associated with heart failure, which has been previously evidenced by clinical outcome data. Overall, MAGEPRO is a powerful tool to enhance inference of gene regulatory effects in underpowered datasets and has improved our understanding of population-specific and shared genetic effects on complex traits.

## Introduction

Many genetic variants drive complex traits by regulating gene expression^1–8^. Confident characterization of genetic effects on gene expression is required for the functional interpretation of disease-associated variants from genome-wide association studies (GWAS)^9–11^. For example, transcriptome-wide association studies (TWAS) integrate GWAS and gene expression data to enable the identification of gene-disease associations, which can reveal genes underpinning disease susceptibility, nominate candidate biomarkers for clinical use, or propel therapeutic development^12–14^. Despite the potential to unravel the functional mechanisms of diseases, our current understanding of disease-critical genes has been limited by variant-to-gene linking strategies that rely heavily on sample size.

Although there is widespread availability of expression quantitative trait loci (eQTL) summary statistics, such as across different human tissues from the Genotype-Tissue Expression (GTEx)^15^ project or from single cell RNA-sequencing data generated by eQTLGen^16^, datasets from non-European populations are severely limited. Differences in allele frequency, linkage disequilibrium (LD), and potentially causal variants reduce the applicability of genetic models (of gene expression and complex traits alike) trained in European populations to non-European populations^17–21^ and therefore limit the relevance of disease-gene associations detected by European TWAS to other global populations. Therefore, there is an urgent need to more accurately infer which genetic variants regulate gene expression and by how much, specifically in understudied populations. Orthogonal to cross-ancestry fine-mapping of TWAS associations^22^, there also exists an opportunity to prune dense genomic loci with multiple gene-disease associations to effects that are shared across ancestries, as causal genes are expected to be shared across ancestries, more so in fact than causal variants.

Efforts to include diverse groups of individuals in genetic studies have yielded a modest number of publicly available eQTL summary statistics from non-European populations^23–27^. Although the statistical power of the eQTL studies performed in non-European populations remains considerably weaker than that of European studies (6.5- and 2.6-fold difference in sample size between European and African American individuals in GTEx^15^ and the Multi-Ethnic Study of Atherosclerosis (MESA)^24^, respectively), these data provide a unique opportunity to capture varying genetic effects on gene expression across diverse ancestries. However, current gene expression prediction models (such as LASSO, elastic net, and the best linear unbiased predictor (BLUP) used in TWAS) can only model the limited individual-level genotype and gene expression data from a single population to compute noisy estimates of variant-gene effect sizes. Previous studies have proven the feasibility of leveraging data from multiple populations to enhance GWAS association power^28^, polygenic risk score (PRS) accuracy^29–31^ and GWAS fine-mapping^32,33^. Thus, we hypothesized that multi-ancestry data would enhance the construction of *cis*-genetic models of gene expression by improving the estimation of variant-level effects and overall expression prediction accuracy. Current multi-ancestry TWAS approaches do not tackle the issue of large uncertainty of inferred *cis*-genetic effects on gene expression in small non-European cohorts. For example, TESLA improves association power by colocalizing a single eQTL dataset with a cross-population meta-analysis of GWAS summary statistics, producing results with mixed or uncertain relevance to each ancestry^34^. Another approach called METRO models the uncertainty of gene expression models across multiple cohorts to maximize colocalization with GWAS^35^, resulting in findings that are highly driven by European data when other gene models are derived from smaller non-European datasets. To date, multi-ancestry data has not been used to reduce uncertainty and improve accuracy of population-specific genetic models of gene expression.

Here, we introduce a new method, Multi-Ancestry Gene Expression Prediction Regularized Optimization (MAGEPRO), that improves gene expression prediction accuracy in underpowered ancestries or undersampled tissues by optimally combining eQTL summary statistics from ancestrally and functionally diverse datasets. We evaluate the robustness of our method in various simulated genetic architectures and compare the predictive performance of MAGEPRO to alternative methods of gene expression prediction, including an adaptation of a multi-ancestry complex trait PRS method called PRS-CSx^30^, using 8 different eQTL cohorts representing 3 ancestries. We additionally applied MAGEPRO gene models to perform TWAS with 15 blood-cell traits and 7 immune-mediated diseases, each represented by GWAS cohorts of individuals of African, European, and Hispanic ancestries, to identify novel disease-gene associations and interrogate the population-specificity of these putative disease genes.

## Results

### Overview of MAGEPRO

MAGEPRO maximizes our ability to infer gene regulatory effects in small sample size eQTL datasets and constructs robust *cis*-genetic models of gene expression that are specific to an ancestry. Given individual-level genotype and gene expression data of the target cohort and external eQTL data from diverse ancestries and tissues, MAGEPRO first estimates effect sizes for single nucleotide polymorphism (SNP)-gene pairs in *cis* that are specific to the target population via a LASSO (L1 norm)-regularized linear regression (**Figure 1**, green box). This step constitutes the conventional TWAS gene expression prediction model. Next, MAGEPRO applies the Sum of Single Effects (SuSiE)^36,37^ regression model to each set of external eQTL summary statistics to identify putative causal variants and estimate posterior effect size estimates for all *cis*-variants (**Figure 1**, blue box). Assuming most causal variants are shared, this step is critical to maximizing the cross-population transferability of information from external datasets to the target cohort. Causal variants are more likely to possess predictive power in the target population compared to variants that merely tag the causal variant; specifically, the causal variant may not be sufficiently tagged in the target population if there are differences in linkage disequilibrium and allele frequency between training and target populations. Finally, our approach finds an optimal ridge (L2 norm)-regularized linear combination of posterior effect size estimates from SuSiE and the target population SNP-gene weights to produce the final gene expression prediction model (**Figure 1**, white box). By utilizing existing fine-mapping frameworks and regularizing the combination of SNP-gene weights across datasets, MAGEPRO is designed to include only information that is potentially relevant to the target population, as opposed to other strategies such as METRO (see above) or a meta-analysis approaches where inferred effect sizes are driven by the largest (European) datasets in the analysis.

**Figure 1.**
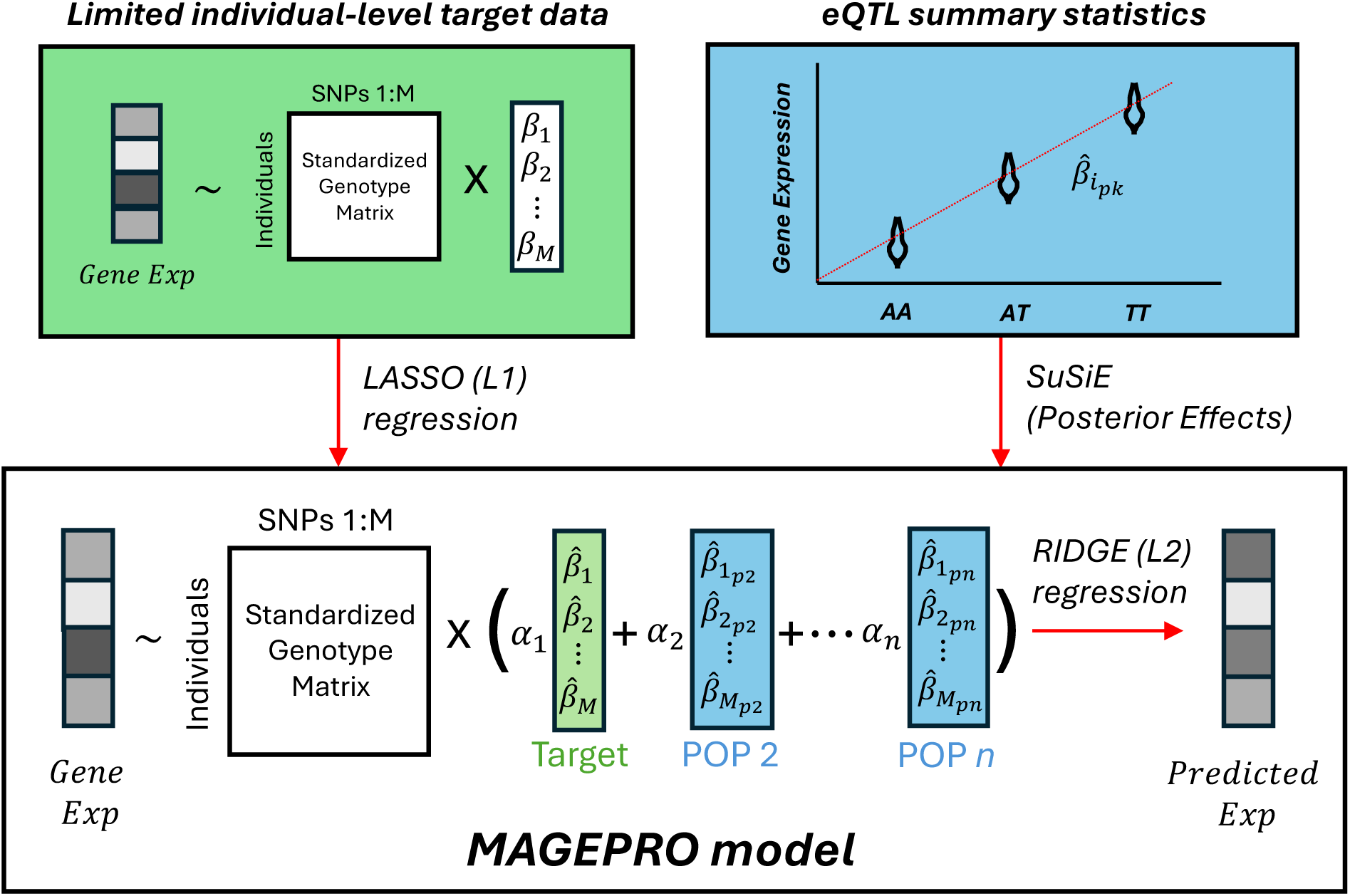
Overview of the MAGEPRO model. Schema of the MAGEPRO model for one gene. MAGEPRO takes limited individual-level target data (green) and external eQTL summary statistics (blue) as input. Red arrows indicate the three main operations of MAGEPRO. First, individual-level gene expression and standardized genotypes are used to estimate noisy effect sizes for the target population (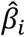 for SNP *i*) using an L1-regularized linear regression. Next, we estimate the posterior effect size estimates for each set of external eQTL summary statistics using SuSiE, designated by 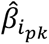 for SNP *i* and population *k*. Finally, we estimate optimal mixing weights of effect sizes across all populations, including the target, using L2-regularized linear regression (*α*_*k*_ for population *k*). The *cis*-heritability of the gene expression 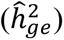 is estimated using the limited individual-level target data and is used to normalize the prediction accuracy 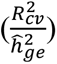 to allow comparisons across genes with different heritabilities.

Throughout this study, we compare MAGEPRO to several methods for gene expression prediction. These include single-ancestry methods commonly used in TWAS, such as LASSO regression^12,14,38,39^, and multi-ancestry approaches, such as a cross-population meta-analysis of eQTL summary statistics. We also utilized methods that are conventionally applied to gene expression or GWAS data, like SuSiE^36,37^ and pruning and threshold (P+T)^17,40,41^. Notably, we benchmarked our tool against a variation of MAGEPRO that we refer to as Multipop, which does not use SuSiE, but rather fits a ridge (L2 norm)-regularized linear combination of raw eQTL summary statistics. Lastly, we benchmarked MAGEPRO against PRS-CSx^30^, a state-of-the-art multi-ancestry PRS method for genome-wide complex trait/disease data. PRS-CSx is a Bayesian framework that models LD heterogeneity across datasets and infers a shared shrinkage parameter to enforce sparsity, which assumes that causal effects are shared, a common assumption of most multi-ancestry fine-mapping models^32,42^. While PRS-CSx is a popular choice for PRS using ancestrally diverse GWAS data^43–49^, this method has not yet been applied to integrate cross-population eQTL summary statistics to create more predictive models of gene expression. In our study, we compare gene expression prediction accuracy 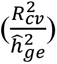 between methods, which is defined as the fraction of gene expression variance explained by the model in cross-validation (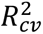), normalized by the upper limit of the prediction: the *cis*-heritability estimated by GCTA^50^ 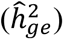. Each competing method is described in further detail in Methods.

### Simulations

We performed extensive simulations to compare the performance of MAGEPRO to the most popular approaches, LASSO for single-ancestry and PRS-CSx for multi-ancestry, under various genetic architectures, using code adapted from the Mancuso Lab TWAS simulator (Code Availability)^51,52^. We used real genotypes from the 1000 Genomes Project^18^ as LD reference panels to simulate genotypes and *cis*-regulated gene expression data across African, European, and American ancestries (Methods). We compared the 5-fold cross-validation accuracy of each model in predicting *cis*-regulated gene expression in African individuals (target), using simulated European and American summary statistics (external) for both PRS-CSx and MAGEPRO. In our primary analysis, we simulated genes with four causal *cis*-eQTLs shared across populations with correlated true effect sizes (r = 0.8); we varied target population sample sizes, the heritability of gene expression, and the number of causal *cis*-eQTLs. In secondary analyses, we varied whether or not eQTL effects were correlated across ancestries, changed whether or not there were ancestry-specific causal *cis*-eQTLs in high LD with the causal variant of the target ancestry, and lastly, evaluated if MAGEPRO can still improve the accuracy of gene models when SuSiE fails to identify a likely causal variant. More details on our simulation framework are described in Methods and the Supplementary Note.

Within our primary analyses, we first compared the prediction accuracy of the three methods, calculated as 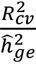 (see above) across target population sample sizes ranging from 80 to 500 individuals and gene expression heritability ranging from 5% to 40%. Across 1,000 independently simulated genes, MAGEPRO outperformed both LASSO and PRS-CSx in each of 20 different sample size and *cis*-heritability settings with an average improvement of 5.7% and 4.5% in accuracy, respectively (**Figure 2A**, **Supplementary Tables 1-2**). Generally, larger sample sizes of the target population resulted in more accurate predictions for a given heritability; and, accuracy notably increased and began to approach 100% for each method within the most heritable genes (40%), thanks to the larger and more easily identifiable eQTL effects. The utility of MAGEPRO is most clearly demonstrated at smaller sample sizes and higher gene expression heritability (**Supplementary Figures 1-2**), enhancing accuracy by > 9% compared to LASSO (*p* < 1.4 × 10^-56^) and by > 7% compared to PRS-CSx (*p* < 2.3 × 10^-49^) when the sample size of the target cohort is 80 individuals and the heritability of the gene is ≥ 20%. For lowly heritable genes, MAGEPRO demonstrates an increasing margin of advantage over the other two methods as sample sizes grow (**Supplementary Figures 1-2**), suggesting that MAGEPRO may be especially useful for modeling the genetic architecture of disease-critical genes whose regulatory effects are flattened by natural selection and thus have lower *cis*-heritability^53,54^.

**Figure 2.**
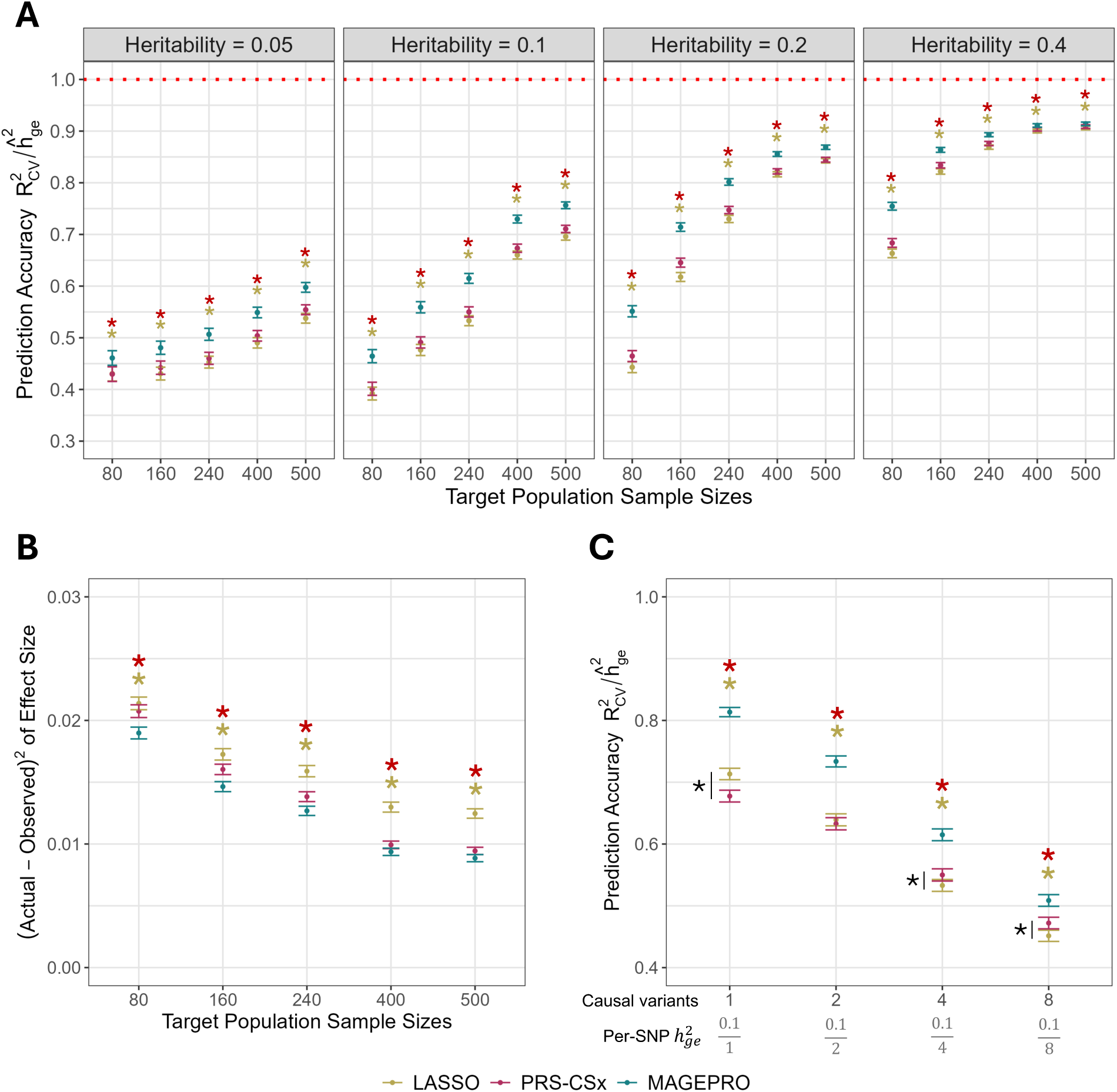
MAGEPRO outperforms alternative gene expression prediction models in various simulated architectures. (A) Predictive accuracy of LASSO, PRS-CSx, and MAGEPRO across different gene expression heritability and sample size settings. Across all settings, genes were simulated with four causal variants. Accuracy is calculated as the ratio of the cross-validation 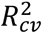 and the GCTA-estimated *cis*-heritability of gene expression 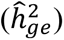. (B) Squared difference between the simulated (actual) and estimated effect sizes of the four causal variants per gene. *Cis*-heritability was set to 10%. (C) Predictive accuracy 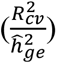 of methods while varying the number of causal variants and maintaining the total *cis*-heritability 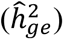 at 10%. Sample size was set to 240. In all panels, data are presented as mean values across 1,000 independently simulated genes with confidence intervals representing ± 1 standard error. Yellow (resp. red) asterisks indicate that the difference between MAGEPRO and LASSO (resp. PRS-CSx) results is significant. Black asterisks highlight pairwise comparisons. All hypothesis tests are two-sided paired t-tests. Numerical results are reported in **Supplementary Tables 1-6**.

We further hypothesized that MAGEPRO would achieve superior prediction accuracy by estimating more accurate eQTL effect sizes. Indeed, when we compare the squared difference between simulated (true) and estimated causal eQTL effect sizes, MAGEPRO produces smaller errors compared to both competing methods across the five different sample sizes at 10% gene expression heritability (all *p* < 3.98 × 10^-4^, **Figure 2B**, **Supplementary Tables 3-4**). Although the accuracy of causal eQTL effect sizes is not a requirement for prediction methods (e.g., prediction can be achieved with strong tagging variants), we believe this characteristic of MAGEPRO may lead to more accurate results from downstream gene-based association analysis like TWAS.

We also evaluated each method across genetic architectures with varying numbers of causal *cis*-eQTLs while maintaining a constant 10% *cis*-heritability and target sample size of 240, which is synonymous with decreasing the per-SNP heritability 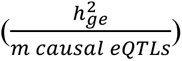. Overall, as the per-SNP heritability decreases, the prediction accuracy of all methods decreases due to the difficulty of capturing larger quantities of smaller effects (**Figure 2C**, **Supplementary Tables 5-6**), exemplifying the challenge of modeling the genetic regulation of disease-critical genes, which are more likely to have lower *cis*-heritability (see above). Despite this challenge, MAGEPRO outperformed both LASSO and PRS-CSx in each per-SNP heritability setting (all *p* < 1.5 × 10^-5^), while PRS-CSx notably surpassed the accuracy of LASSO for the two lower per-SNP heritability settings. This indicates that at current eQTL study sample sizes, leveraging multi-ancestry data is a useful tool for accurately modeling the genetic regulation of potentially disease-relevant genes and may help more confidently identify which diseases they influence via gene-based association tests.

In secondary analyses, we tested the performance of MAGEPRO when the effect sizes of shared causal *cis*-eQTLs are drawn independently across ancestries and are thus uncorrelated. Although MAGEPRO achieves larger improvements relative to LASSO and PRS-CSx when effect sizes are correlated across ancestries, our tool robustly improves prediction accuracy even when effect sizes are independent (**Supplementary Figure 3**) and trends across sample sizes and heritabilities are largely shared with simulations with correlated eQTL effects. Recent work shows that effect size correlations across ancestries are lower for loss-of-function intolerant genes^39^ and variants with ancestry-specific disease effects may reside closer to genes interacting with the environment, such as immune responses^55^. This suggests that MAGEPRO will continue to improve gene model accuracy, even when causal eQTL effect sizes are independent, which could potentially lead to the discovery of novel gene-disease associations. In a related framework, we simulated gene expression prediction models based on a single causal eQTL in the target African population. In this analysis, the single causal eQTL is not shared across any ancestries, but the two causal variants from the European and American populations are in high LD with the causal variant of the target population (**Supplementary Figure 4**). Overall, we observed highly similar trends with that of **Figure 2**; in fact, the accuracies across sample sizes and heritabilities were greater than in **Figure 2** due to the fact that per-SNP heritability was proportionally higher thanks to simulating a single causal variant.

Lastly, we explored whether the improvement in accuracy provided by MAGEPRO depends on the ability of SuSiE to identify causal *cis*-eQTLs in external datasets. The enhancement of prediction accuracy relative to LASSO is nominally larger when SuSiE identifies at least 1 causal *cis*-eQTL (*PIP* ≥ 0.95) across the external datasets and this difference is only statistically significant at the largest target population sample size of 500 (*p* = 0.03) (**Supplementary Figure 5**). This implies that although isolating the causal regulatory variants contributes to improved prediction, MAGEPRO does not rely on fine-mapped SNPs with high PIPs, but rather on posterior effect size estimates.

### Benchmarking MAGEPRO against alternative gene expression prediction methods

In real data analysis, we employed MAGEPRO to create *cis*-genetic models of gene expression for 8 eQTL cohorts across 3 different ancestries (average *n* = 355) using up to 5 external summary statistic datasets as features in the MAGEPRO model (**Table 1**)^15,16,24,25,27^. For each gene, we performed variable selection, e.g., eQTL fine-mapping, applying SuSiE to each summary statistic dataset (Methods). We explored the possibility of leveraging IMPACT, a tool we have previously developed to estimate the probability that a variant participates in cell-type-specific gene regulation^56^, as Bayesian SNP-selection priors in SuSiE have been shown to improve fine-mapping power^57^. Although this increased the number of genes with at least 1 putatively causal eQTL (posterior inclusion probability (PIP) ≥ 0.95), increased average PIPs in credible sets, and decreased average credible set size, it did not substantially affect the accuracy of MAGEPRO gene models (**Supplementary Figures 6-7**). Even random priors seemed to improve fine-mapping metrics, likely by randomly pruning high PIP variants in high LD; but, ultimately the predictive capacity of posterior effect size estimates do not strictly depend on reduced credible set size and high PIP SNPs, thus the gene model accuracy is not necessarily affected (**Supplementary Figure 6**). These results are consistent with our simulations that indicated MAGEPRO need not find a putatively causal eQTL to enhance prediction accuracy relative to LASSO. Therefore, we elected to not use IMPACT priors in the default implementation of MAGEPRO.

**Table 1.**
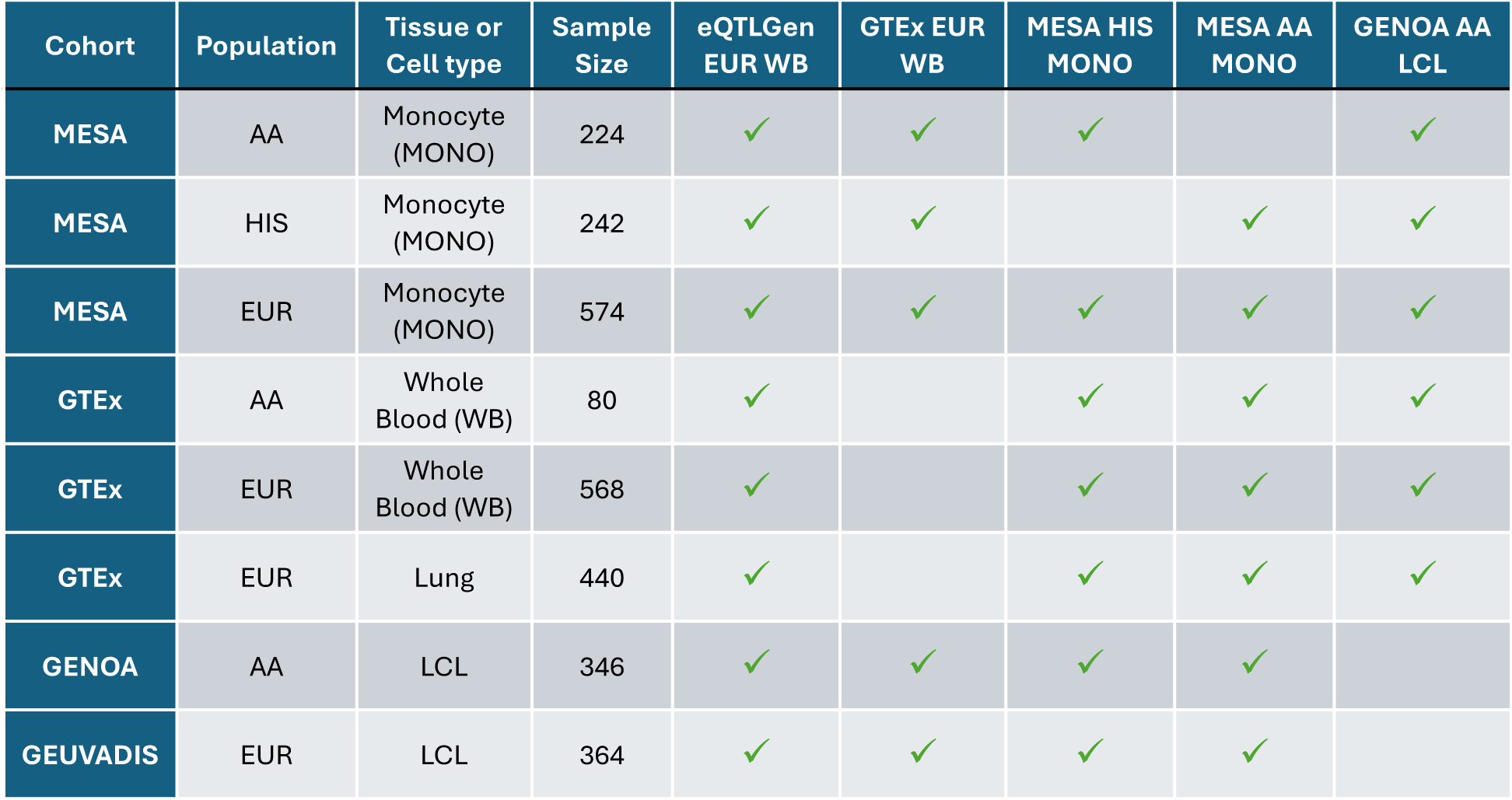
External eQTL summary statistics used for each target cohort. Rows correspond to each target cohort for which individual-level gene expression and genotype data were used to create genetic models of gene expression. The last five columns correspond to external eQTL summary statistics used as inputs to MAGEPRO. We avoided using external summary statistics that contain the same individuals as the target cohort to prevent over-fitting and inflation of cross-validation results. Sample sizes indicate the number of individuals in a target cohort after relatedness-based filtering (Methods). AA, African American; HIS, Hispanic/Latino; EUR, European; LCL: lymphoblastoid cell line.

Next, we applied GCTA to each target eQTL cohort to estimate the *cis*-heritability 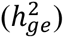 of each gene. For genes with larger *cis*-heritability estimates, SuSiE detected a larger number of putatively causal eQTLs on average (*PIP* ≥ 0.95) (**Supplementary Figure 8**). We also observed that the estimated *cis*-heritabilities of gene expression were highly correlated across ancestries, consistent with previous work^22^ (Pearson correlation (r) ranging from 0.32 to 0.83 in comparisons between European, Hispanic/Latino, and African American populations) (**Supplementary Figure 9**). However, we observed similar heterogeneity of heritability estimates even across cohorts within the same ancestry (r = 0.34, 95% CI [0.311, 0.367]) between European individuals in GEUVADIS and GENOA cohorts), suggesting that cross-cohort variation may limit out-of-cohort prediction accuracy.

We next compared the performance of various methods in predicting expression levels of significantly *cis*-heritable genes in each target cohort 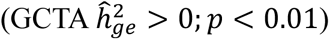. These methods, introduced above and in more detail in Methods, comprise a cross-population meta-analysis, pruning and thresholding (P+T) of target marginal *cis*-eQTL, LASSO of the target population, SuSiE applied to the target population, a ridge (L2 norm) regression of full external *cis*-eQTL summary statistics (which we refer to as “Multipop”), PRS-CSx, and MAGEPRO. We note that not all external summary statistics contain associations for all genes, and thus MAGEPRO utilizes only relevant external datasets available to each gene.

First, we applied each method to predict lymphoblastoid cell line (LCL) gene expression in the Genetic Epidemiology Network of Arteriopathy (GENOA) African American (AA) cohort (*n* = 346). MAGEPRO outperformed all competing methods (all paired one sided t-test *p* < 3 × 10^-10^) and improved prediction accuracy by 10.4% relative to LASSO averaged across 4,141 *cis*-heritable genes (**Figure 3A**, **Supplementary Table 7**). MAGEPRO’s accuracy exceeded that of Multipop (*p* = 8 × 10^-32^), suggesting that the posterior effect sizes estimated by SuSiE are prioritizing variants that are critical in predicting gene expression. Notably, our model increased prediction accuracy relative to LASSO by over 20% for 1,177 genes and introduced 204 new genes with an 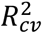 significantly greater than 0 (*p* < 0.05). We then down-sampled the GENOA AA cohort (*n* = 100) to challenge MAGEPRO in a small sample size setting (one that is similar to the number of African American individuals in GTEx). We found that MAGEPRO maintains improved accuracy compared to all methods when target population genotype and gene expression data is extremely limited (*p* < 0.01 across all comparisons, **Figure 3B**, **Supplementary Table 8**). At this sample size, we achieved a 4.4% improvement in accuracy relative to PRS-CSx (*p* = 4 × 10^-10^), suggesting that the layers of regularization in our framework minimize overfitting even with small training cohorts.

**Figure 3.**
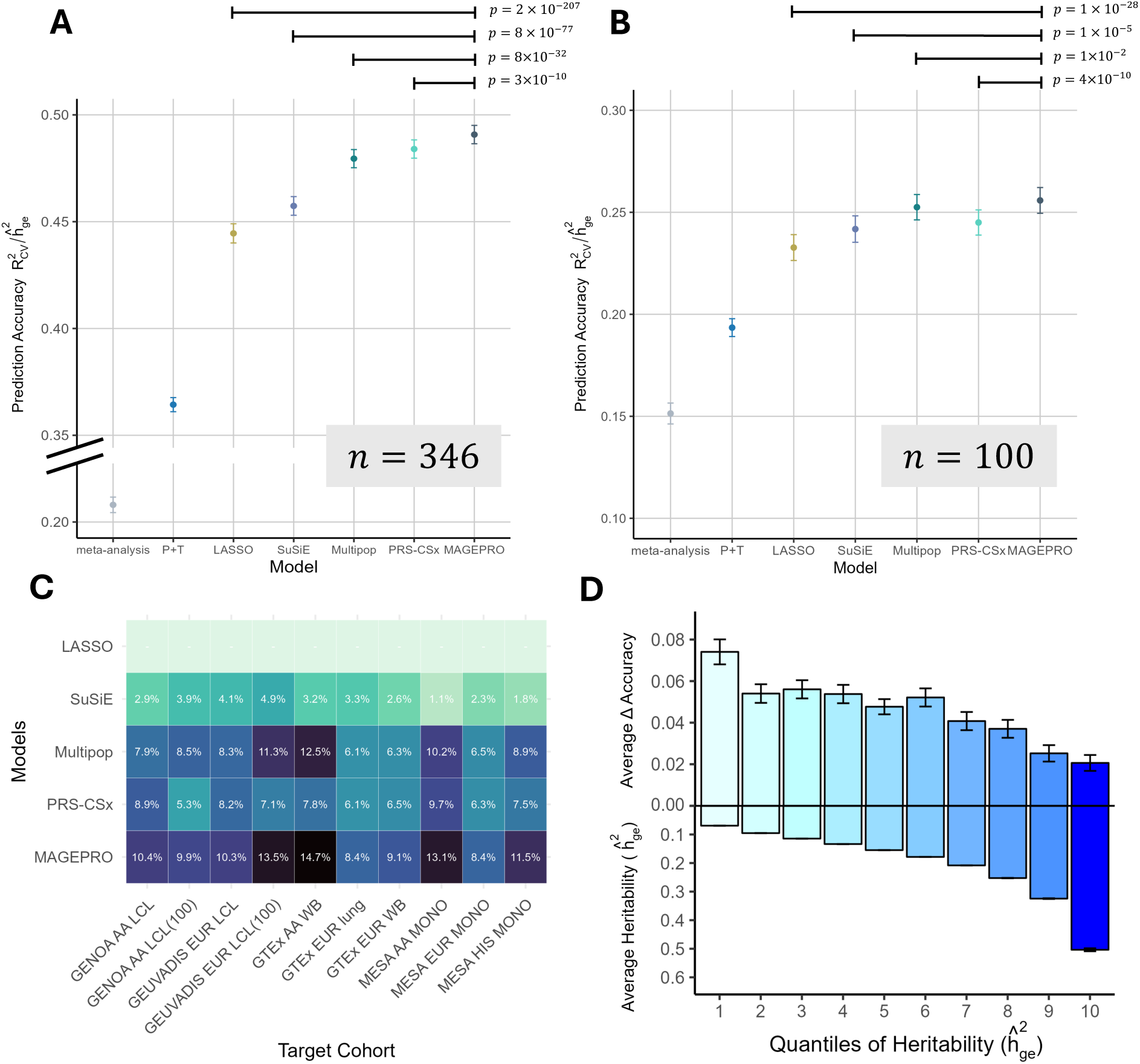
MAGEPRO outperforms alternative methods in real data. (A,B) Comparison of the accuracy of different models in predicting LCL gene expression in the GENOA AA population at two different sample sizes (A: full cohort, B: random down-sampling to 100 individuals). P-values are derived from a one-sided paired t-test, testing the alternative hypothesis that MAGEPRO produces larger accuracies. Comparisons between MAGEPRO and meta-analysis or P+T not annotated due to low precision to estimate such small p-values. (C) Performance of the top five gene expression prediction methods across the eight different target cohorts from **Table 1** plus two randomly down-sampled cohorts, indicated by (100). Values in the heatmap are the percent change in predictive accuracy relative to LASSO regression. All percent differences are significant according to one-sided paired t-tests (p < 0.05). (D) The average change in accuracy between MAGEPRO and LASSO 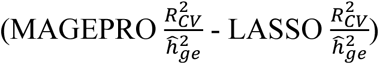 across 10 quantiles of genes grouped by GCTA-estimated *cis*-heritability. Accuracy and heritability values were estimated for LCL gene expression in the GENOA AA population. In panels A, B, and D, data are presented as mean values with confidence intervals representing ± 1 standard error. LCL, lymphoblastoid cell line; AA, African American. Numerical results are reported in **Supplementary Tables 7-10**.

We observed similar trends across all 8 target eQTL cohorts (10 including down-sampled cohorts). In predicting monocyte gene expression in the Multi-Ethnic Study of Atherosclerosis (MESA) Hispanic/Latino (HIS) cohort, MAGEPRO again outperformed all competing methods (all *p* < 6 × 10^-21^), improving prediction accuracy relative to LASSO by over 20% for 942 genes and creating 191 new gene models with significantly positive 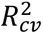 (**Supplementary Figure 10**). MAGEPRO improved prediction accuracy relative to LASSO by 14.7% in the GTEx AA cohort (*n* = 80, Whole Blood) and by 13.5% in the down-sampled GEUVADIS European (EUR) cohort (*n* = 100, LCL), suggesting that our method provides the largest relative improvement when the target cohort sample size is limited (**Figure 3C**, **Supplementary Table 9**).

Next, we aimed to characterize the genes for which MAGEPRO is most useful for capturing the *cis*-genetic component of expression. We observed that the change in accuracy between MAGEPRO and LASSO 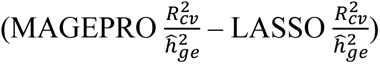 is negatively correlated with *cis*-heritability estimates (*r* = −0.14, *p* = 3.7 × 10^-29^ and *r* = −0.17, *p* = 4.7 × 10^-23^ for GENOA AA and MESA HIS respectively; **Figure 3D**, **Supplementary Table 10**, **Supplementary Figure 11**). This indicates that MAGEPRO offers the greatest modeling improvements to low heritability genes, which are more likely to be disease-critical, as natural selection restricts the magnitude of *cis*-genetic effects (and thus heritability) on disease-critical genes. For example, we found that loss-of-function intolerant genes (pLI > 0.9)^58^ indeed have the lowest gene expression heritability estimates (**Supplementary Figure 12**, *p* < 7.0 × 10^-8^). Additionally, we found that MAGEPRO offers the greatest advantage over PRS-CSx when the per-SNP heritability of the gene 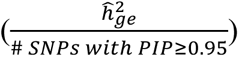, which is proportional to the power to detect *cis*-genetic effects^31^, is low (**Supplementary Figure 13**).

We also evaluated the generalizability of each model to individuals from a different study cohort in the same target ancestry. To this end, we compared out-of-cohort prediction accuracy. We trained gene expression prediction models in GENOA AA and GEUVADIS EUR cohorts, each at two different sample sizes, and then applied these models to predict LCL gene expression in GEUVADIS Yoruba (YRI) and GENOA EUR cohorts, respectively. MAGEPRO and SuSiE consistently outperformed the other methods (LASSO, Multipop, PRS-CSx) in out-of-cohort prediction, suggesting that frameworks which prioritize putative causal eQTL may result in more generalizable predictive models (**Supplementary Figure 14**). We note that we did not assess cross-population meta-analysis or P+T in this analysis, as they performed much more poorly in within-cohort cross-validation tasks. However, the performance of MAGEPRO relative to SuSiE (applied directly to the training population) was highly variable. For example, the SuSiE model trained in the down-sampled GENOA AA cohort (*n* = 100) achieved a higher out-of-cohort *R*^2^ than MAGEPRO (*p* = 0.006, **Supplementary Figure 14**), possibly due to the different extent of admixture between African American (training) and Yoruba individuals (testing) (**Supplementary Figure 15**) or due to the inherent cross-cohort variation in the genetic architecture of gene expression that we previously observed (**Supplementary Figure 9**). In contrast, the MAGEPRO model trained in the down-sampled GEUVADIS EUR cohort (*n* = 100) exceeded SuSiE in out-of-cohort prediction by 6% (*p* = 8.6 × 10^-9^, **Supplementary Figure 14**). MAGEPRO generally excels in out-of-cohort prediction when the genetic ancestry of the training and testing cohorts are closely related (**Supplementary Figures 14-15**), highlighting the population-specific nature of MAGEPRO models. In other words, SuSiE applied to the target training population is effective at assaying causal variants that are likely to be shared across populations, but more population-specific effects may be identified by MAGEPRO, which is tailored to the training population.

We found that MAGEPRO is consistently most useful when the target population genotype and gene expression data is limited. We hypothesized that this may include situations where the target tissue is less accessible and/or data is scarce. Therefore, we explored if genetic models of gene expression in tissues that are seemingly unrelated to blood can be improved by integrating widely available blood-derived eQTL summary statistics. To this end, we applied MAGEPRO to create Lung gene models in GTEx using blood-related external *cis*-eQTL summary statistics (**Table 1**). MAGEPRO produced impressively accurate gene models (59% on average) while outperforming all competing methods (all *p* < 1 × 10^-46^), likely owing to the correlation of *cis*-genetic regulation of gene expression across tissues^15^ (**Supplementary Figure 16**), not unlike the cross-population sharing of causal effects. Moreover, this suggests that MAGEPRO successfully identifies regulatory effects from blood tissue that are transferable to lung tissue, notably resulting in an 8.4% average improvement over the lung-specific LASSO model (*p* = 9 × 10^-307^).

We implemented MAGEPRO as a publicly available pipeline on GitHub (Code Availability), leveraging multiple threads on both high-performance computing (HPC) clusters^59^ and personal devices to enhance computational efficiency (**Supplementary Figure 17**).

### Transcriptome-wide association studies are sensitive to cis-genetic models of gene expression

We hypothesized that one of the most compelling applications of MAGEPRO would be to make the inference of disease-critical genes more powerful for underrepresented populations. To this end, we applied LASSO, SuSiE, PRS-CSx, and MAGEPRO models trained in 7 blood-related eQTL cohorts (MESA AA Monocyte, GENOA AA LCL, GTEx AA Whole Blood, MESA EUR Monocyte, GEUVADIS EUR LCL, GTEx EUR Whole Blood, MESA HIS Monocyte) to perform TWAS for 15 blood cell traits and 7 immune-mediated diseases using ancestry matched GWAS summary statistics from Chen and colleagues^60^ (AFR *N* = 13,391, EUR *N* = 516,979, HIS *N* = 6,849) and the Global Biobank Meta-analysis Initiative (GBMI)^61^ (AFR *N* = 26,052, EUR *N* = 1,024,298, Native American ancestry (AMR) *N* = 15,490), respectively (**Supplementary Table 11**). We note that we did not have access to AMR eQTL data and, therefore, we used HIS gene expression prediction models as proxies to perform TWAS in the AMR population. To avoid complicated notation, we refer to subsequent TWAS analysis involving HIS eQTL data and AMR GWAS as HIS. Generally, we observed two main phenomena. In one case, MAGEPRO models led to more accurate *cis*-genetic models of gene expression (relative to LASSO), and this subsequently eliminated the statistically significant TWAS association observed for LASSO. In the other case, MAGEPRO generated predictive gene expression models (significantly positive R^2^) even though LASSO failed to do so; this resulted in many new gene-trait/disease associations, exemplifying the utility of MAGEPRO to enhance disease inference in underpowered cohorts and underrepresented populations. Ultimately, both of these scenarios allowed us to explore the sensitivity of TWAS to slight variations in *cis*-genetic gene models. We explore examples of both cases below in more depth.

First, we observed that the average change in gene expression prediction *R*^2^ (MAGEPRO *R*^2^ – LASSO *R*^2^) does not correlate with the average change in TWAS chi-square statistic (χ^2^) (MAGEPRO TWAS χ^2^ - LASSO TWAS χ^2^) across significantly *cis*-heritable genes (**Supplementary Figure 18**). This result is not surprising as few genes play critical roles for any one disease, and MAGEPRO is able to improve the mapping of *cis*-genetic effects for both disease-critical and non-critical genes. However, this observation led us to understand that sometimes an improved gene expression prediction model may actually produce a weaker TWAS association, implying that less accurate gene models were only spuriously correlated with disease. In other words, MAGEPRO provides an additional utility of enhancing the confidence in TWAS association results by increasing the gene expression prediction accuracy. While TWAS is most well-powered to identify genes with large *cis*-genetic effects that colocalize with disease, our observation here does not invalidate the compelling nature of our previous finding that MAGEPRO produces the largest improvements in model accuracy for low heritability genes, which due to natural selection may be more disease-critical. Therefore, by learning more accurate *cis*-genetic models of gene expression, MAGEPRO may be additionally poised to help derive disease-critical effects on gene expression in frameworks beyond TWAS.

There were several genes for which the conventional single population TWAS model produced a significant TWAS association that was ablated when the gene model was improved with MAGEPRO. For example, the association between *ZNF213-AS1* and red blood cell count in the African American population diminished as MAGEPRO improved the accuracy of gene expression prediction (**Figure 4A**, **Supplementary Table 12**). Investigating how the *cis*-genetic model of gene expression colocalizes with GWAS summary statistics reveals that the MAGEPRO model captured a new eQTL signal (“MAGEPRO-specific” in teal), improving gene expression prediction accuracy (from 24% with SuSiE or 33% with LASSO to 45% with MAGEPRO) but providing conflicting evidence against the negative association with the GWAS phenotype (**Figure 4A**). *ZNF213-AS1* is a noncoding antisense RNA gene which controls breast cancer progression by modulating estrogen receptor signaling^62,63^, but links to blood-related phenotypes have not been reported in the literature. Additionally, this association was not found in the European TWAS (z = −2.8, not significant [n.s.]), although the gene model achieved near perfect accuracy. To summarize, while TWAS does not account for the uncertainty of gene expression models, our findings suggest that considering association statistics across different models for the same gene can reveal unstable gene-disease associations and potentially false positives.

**Figure 4.**
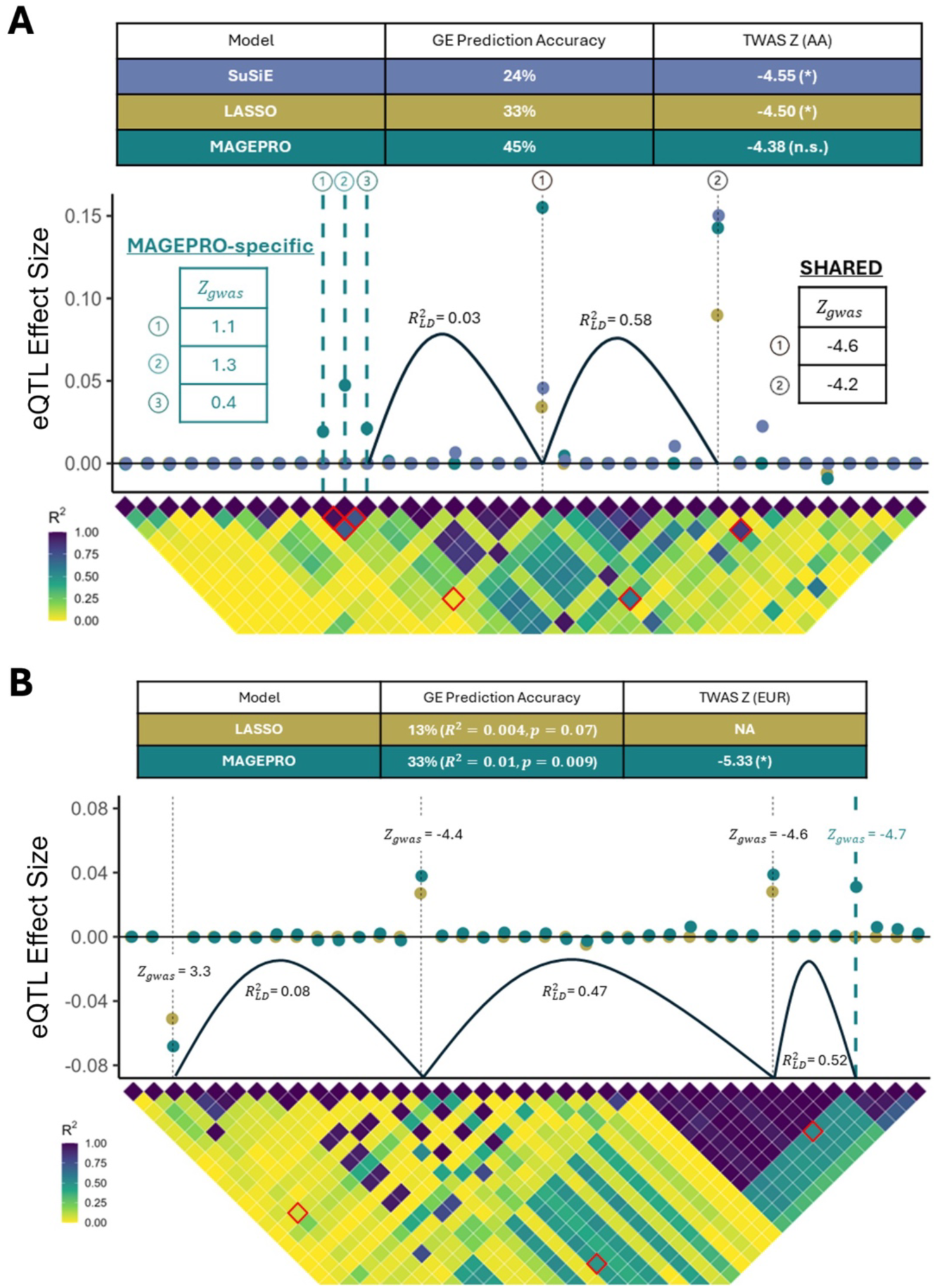
Gene-disease associations are sensitive to gene expression prediction models. (A) An example of a TWAS association that becomes non-significant after MAGEPRO has improved the accuracy of the gene model. MAGEPRO introduces three variants to the gene model for *ZNF213-AS1,* trained on GENOA LCL data from African American individuals, that do not colocalize well with a GWAS for red blood cell count (teal). (B) An example of a new TWAS association introduced by MAGEPRO. *RGS14* is newly associated with asthma based on a European monocyte model from the MESA cohort. In both panels, the asterisk (*) indicates significance using a Bonferroni threshold across cohort-specific *cis*-heritable genes (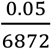 for A, 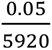 for B). The dot plot shows the effect sizes inferred by the *cis*-genetic model of gene expression created by each method. Black dotted vertical lines designate eQTL effects identified by all/both models and teal dotted vertical lines designate effects captured specifically by MAGEPRO. In the heatmap below, 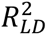 values that are relevant to potential interactions between variants are boxed in red. Distances between SNPs are not to scale; the x-axis indicates the indices of the *cis*-SNPs ordered by increasing genomic coordinate. LCL, lymphoblastoid cell line; GE, gene expression. Numerical results are reported in **Supplementary Tables 12-13**.

Second, we observed that modest changes to *cis*-genetic models of gene expression can also give rise to biologically plausible new disease-gene associations. For instance, *RGS14* was not analyzed in the European TWAS using LASSO because the model produced an *R*^2^ that was not significantly greater than 0 (**Figure 4B**, **Supplementary Table 13**). The MAGEPRO model introduced a new eQTL signal (teal dotted line), which helped the model achieve a significantly positive *R*^2^ (*p* < 0.05) and provided additional evidence to the negative association with asthma (**Figure 4B**). The estimated heritability 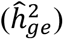 of *RGS14* was only 0.03 (se = 0.015), reflecting the inherent difficulty in modeling genetic effects on genes with low heritability and the utility of MAGEPRO for detecting putative disease-critical genes that could not previously be reliably analyzed. *RGS14* belongs to a family of proteins that regulate G protein signaling, which plays a significant role in asthma^64,65^. Current asthma therapies include G protein signaling agonists and antagonists, which relax airway smooth muscles and reduce airway inflammation, respectively^66^. Our finding suggests that regulatory variants modulating G protein signaling may carry genetic risk for asthma.

### MAGEPRO recapitulates gene-disease associations across diverse ancestries and reveals ancestry-specific findings

Now that we understand the dominant mechanisms by which MAGEPRO can inform gene-disease association studies (e.g., by ablating the significant association producd by less accurate models, or by producing significant associations for genes that previously lacked predictive models), we sought to apply our models across diverse ancestries to characterize population-specific or population-shared gene-level effects on complex traits and diseases. We organized our analysis into two disjoint sets of genes: those with fairly accurate predictive models (*R*^2^ > 0, *p* < 0.05) across all methods (LASSO, SuSiE, PRS-CSx, MAGEPRO) and those that lacked a predictive LASSO model.

We first analyzed all genes with a gene expression prediction *R*^2^ significantly greater than 0 in all methods. Aggregating results across 7 blood-related eQTL cohorts and 66 GWAS summary statistics (accounting for 22 unique diseases/traits and 3 ancestries), MAGEPRO identified 2,521 gene-trait associations 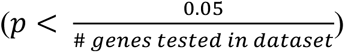 that were not found by LASSO (**Supplementary Table 14**). Considering all four methods, we found that MAGEPRO identified 1,350 significant gene-trait associations that are not identified by any other model (**Supplementary Table 15**), showcasing the benefit of MAGEPRO in augmenting current gene expression prediction models in the TWAS framework. However, MAGEPRO gene models do not necessarily generate more significant gene-trait associations than other methods (**Supplementary Figure 19**). This is because improving genetic models of gene expression yields TWAS results that are more reliable, but not necessarily stronger in association as we discussed previously (**Figure 4A**, **Supplementary Figure 18**). When we applied Monocyte gene models trained in MESA African American individuals to TWAS, MAGEPRO found 8 significant associations not identified by LASSO (6 of them as a result of larger gene model *R*^2^) (**Figure 5A**, **Supplementary Table 16**) and 20 significant associations not found by PRS-CSx (10 of them as a result of larger gene model *R*^2^) (**Figure 5B**, **Supplementary Table 17**). In contrast, when we applied our LCL gene models trained in GENOA African American individuals, PRS-CSx identified 16 associations not found by MAGEPRO (**Supplementary Figure 19**). However, MAGEPRO produced a more accurate genetic model of gene expression for 9 of these 16 genes, suggesting that a majority of the gene-trait associations undetected by MAGEPRO may be false positives, or at the least, unreliable associations. We found similar patterns when comparing TWAS associations across MAGEPRO, LASSO, and PRS-CSx in Hispanic/Latino individuals (**Supplementary Figure 20**), although the limited GWAS sample size for this population greatly reduced our power to assess patterns of gene-trait associations across methods. Reflecting on our results, our suggested best practice is to use the most accurate *cis*-genetic model of gene expression for each gene, as similarly implemented in FUSION. Although it does not always lead to more statistically significant gene-trait associations (**Supplementary Figure 19**), TWAS results will be more credible when the gene expression prediction models are more accurate.

**Figure 5.**
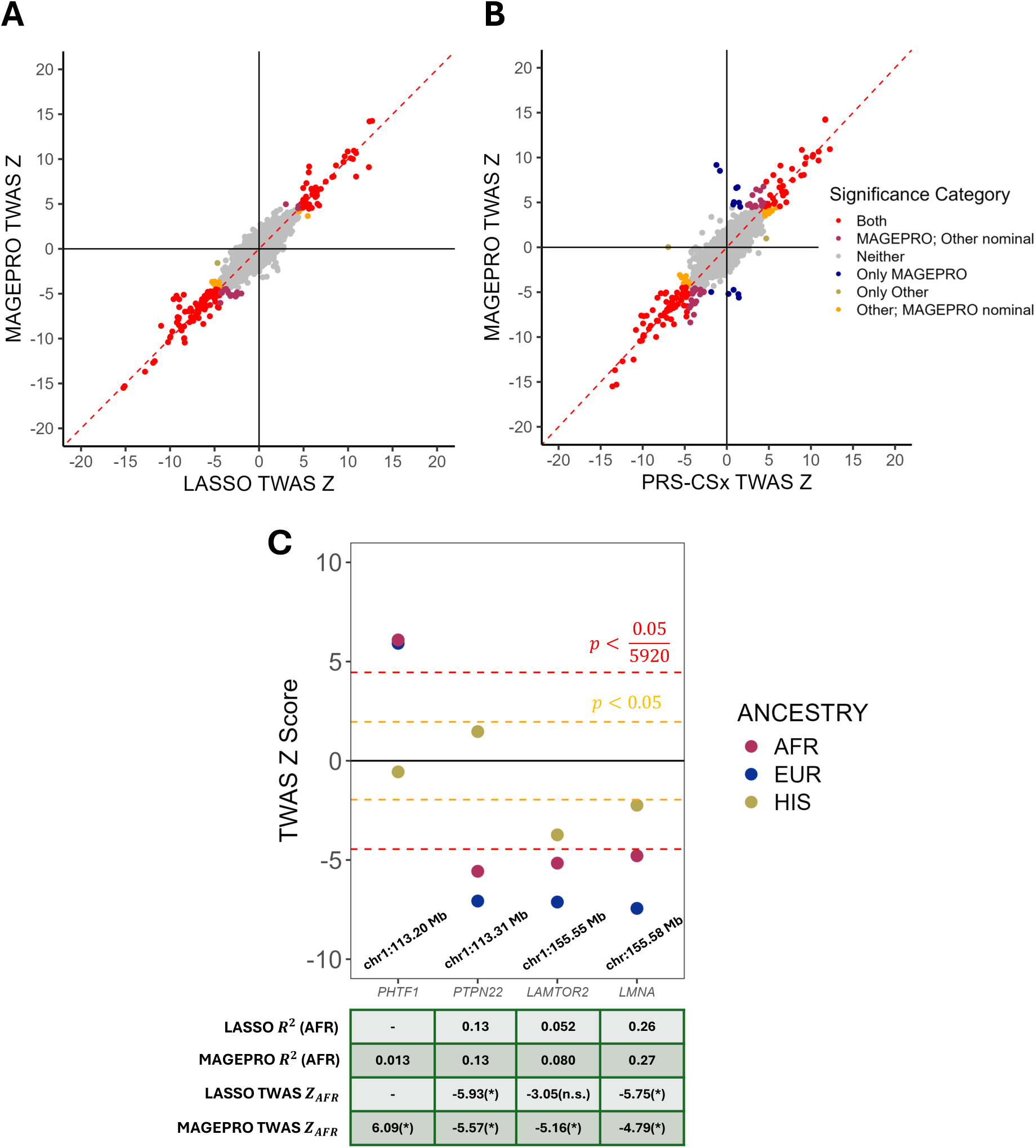
Applying MAGEPRO to improve Monocyte gene expression prediction across three ancestries identifies novel genes associated with blood cell traits. (A,B) Comparison of TWAS z-scores between MAGEPRO and other gene expression prediction methods for the African (AFR) ancestry. Colors correspond to groups of significance described in the legend. “Other” refers to the model in comparison on the x-axis. Results are aggregated across 15 blood cell traits. (C) Miami plot of TWAS associations with white blood cell counts across three different ancestries. Green table display the gene expression prediction *R*^2^ and TWAS z-score in the AFR population (statistics for other population are presented in **Supplementary Figure 22**). Positions indicate the start of the *cis* window, nominal significance threshold is *p* < 0.05 and Bonferroni significance threshold is 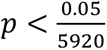 for all panels. Numerical results are reported in Supplementary Tables 16-18.

Second, we explored how improving genetic models of gene expression in underpowered ancestries can help us challenge or recapitulate results from European TWAS studies. To this end, we investigated TWAS results for white blood cell (WBC) count using Monocyte gene models developed for European, African American, and Hispanic populations; we focus on 4 associations that were consistent across at least two populations: *PHTF1, LAMTOR2, PTPN22,* and *LMNA* (**Figure 5C**, **Supplementary Table 18**)*. PHTF1* was not evaluated in African-ancestry TWAS with LASSO because the gene model *R*^2^ was not significantly greater than 0. However, MAGEPRO improved this gene expression prediction model and identified a positive association with WBC count, recapitulating findings from the European population (**Figure 5C**, **Supplementary Figure 21**). *PHTF1* has been associated with other immune-mediated diseases, such as type 1 diabetes in early genetic studies^67^. Additionally, differential expression analysis has shown that this gene is overexpressed in patients with acute lymphoblastic leukemia^68^, a condition characterized by the overproduction of immature white blood cells. This indicates that *PHTF1* is a plausible candidate for regulating white blood cell count and extreme dysregulation of this gene may be linked to forms of leukemia. Furthermore, leveraging MAGEPRO to improve the genetic model of gene expression for *LAMTOR2* by 54% resulted in a new association for individuals of African ancestry, which is consistent with findings from European TWAS (**Figure 5C**, **Supplementary Figure 21**). Previous work shows that experimental knockout of *LAMTOR2* results in an expansion of conventional dentritic cells in mice^69^ and the deficiency of this gene causes immunodeficiency syndromes in humans^70,71^. The replication across ancestries and the layers of evidence in the literature suggest that *LAMTOR2* is another candidate regulator of white blood cell count in humans. *PTPN22,* a well-known regulator of immune signaling^72–76^, and *LMNA*, a major component of the mammalian lamina with important functions in immune cells^77^, was also identified by TWAS for both African and European ancestries using either LASSO or MAGEPRO models. Our findings demonstrate that applying MAGEPRO to improve genetic models of gene expression in understudied populations can help identify potentially causal disease/trait-associated genes that replicate across different ancestries.

Third, we evaluated MAGEPRO’s capacity to identify ancestry-specific gene-trait associations. To achieve this, we analyzed genes with a gene expression prediction *R*^2^ significantly greater than 0 in both LASSO and MAGEPRO and used the better-performing model for TWAS. We identified 137 associations in African or Hispanic populations which were not found in European TWAS (**Supplementary Table 19**). Among these, 13 genes were exclusively identified by MAGEPRO, 5 by LASSO, and 119 by both methods. Notably, MAGEPRO improved the predictive performance of the *UBAP2L* Monocyte gene model in the African American population, modestly raising the *R*^2^ from 0.10 (LASSO) to 0.11. As a result, MAGEPRO detected an association between *UBAP2L* and neutrophil count (NEU) (z = −6.02), which was not found by any European model across monocyte, LCL and whole blood tissues. Previous experimental studies have demonstrated that *UBAP2L* plays a crucial role in the regulation of long-term hematopoietic stem cells^78^, supporting its potential as a candidate regulator of neutrophil counts.

Lastly, we sought to use MAGEPRO to identify disease-critical roles specifically for genes that lacked a predictive LASSO model (*R*^2^ not significantly positive), and thus could not be previously analyzed by TWAS. In this category, MAGEPRO offered 3,195 new gene models across 7 eQTL cohorts. The *cis*-genetic effects of these genes were inherently difficult to model due to the low heritability of gene expression (average 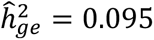, lowest quantile in **Figure 3D**). Nevertheless, MAGEPRO enhanced the average *R*^2^ of these models from 0.0047 with LASSO to 0.031 (a 560% increase). Applying these newly modeled genes to TWAS across all 66 traits yielded 981 associations at Bonferroni significance, where a different threshold was determined for each of 7 eQTL cohorts (**Figure 6**, **Supplementary Table 20**). Several of these associations recaptiulate existings results from colocalization analysis using European GWAS. For example, European MAGEPRO models identified an association of *IRF8*^79^ to monocyte count (MON) and *RCCD1*^80,81^ to red blood cell distribution width (RDW), which are consistent with European colocalization analyses^82,83^ (**Figure 6**). Additionally, some of these associations replicate previously reported European TWAS results in an understudied ancestry. For instance, the relationship between *FAM234* and mean corpuscular hemoglobin concentration (MCHC) has been established in European TWAS^84–86^, but to our knowledge has not been reported using genetic associations from individuals of African ancestry until now (**Figure 6**).

**Figure 6.**
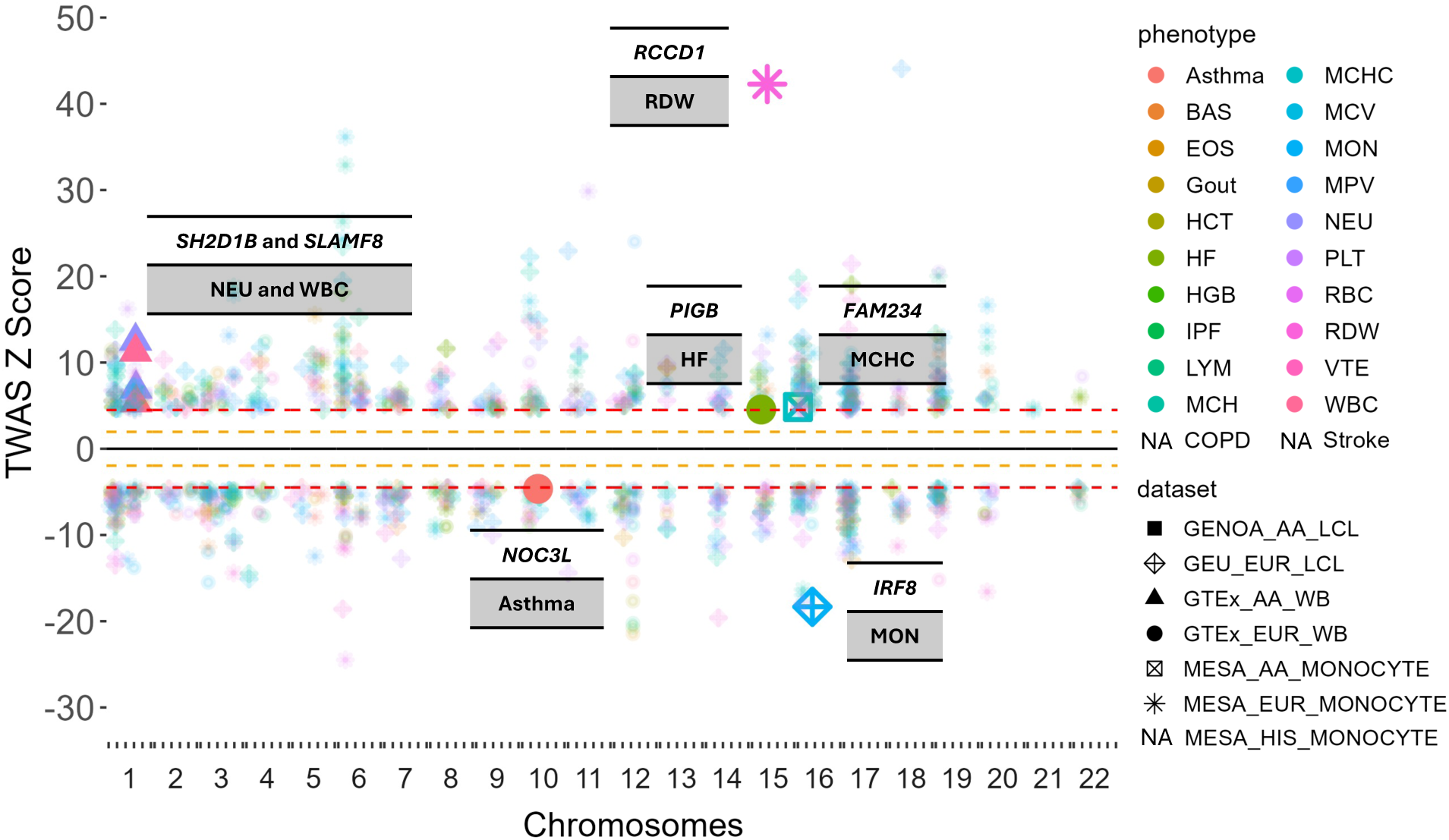
New gene models created by MAGEPRO recapitulate previous findings and identify several biologically plausible new findings. Miami plot shows genome-wide signed TWAS associations from analysis of 22 unique complex traits/diseases across three ancestries (represented by seven independent cohorts). Only gene-trait associations resulting from new gene models created by MAGEPRO (MAGEPRO *R*^2^ > 0, *p* < 0.05 while LASSO *R*^2^ not significantly greater than 0) and passing Bonferroni significance are plotted. Phenotypes and datasets are labeled “NA” if there are no such associations. Examples highlighted in the text are labeled with the gene symbol, associated phenotype, and enlarged point. Yellow dotted line indicates nominal *p* < 0.05 and red dotted line indicates Bonferroni threshold 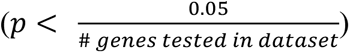. BAS, basophil count; EOS, eosinophil count; HCT, hematocrit; HF, heart failure; HGB, hemoglobin concentration; IPF, idiopathic pulmonary fibrosis; LYM, lymphocyte count; MCH, mean corpuscular hemoglobin; COPD, Chronic obstructive pulmonary disease; MCHC, mean corpuscular hemoglobin concentration; MCV, mean corpuscular volume; MON, monocyte count; MPV, mean platelet volume; NEU, neutrophil count; PLT, platelet count; RBC, red blood cell count; RDW, red blood cell distribution width; VTE, venous thromboembolism; WBC, total white blood cell count. Numerical results are reported in **Supplementary Table 20**.

The new MAGEPRO gene models also resulted in biologically plausible novel findings. For example, African American MAGEPRO models for whole blood identified an association between *SH2D1B* and *SLAMF8* to both neutrophil count (NEU) and white blood cell (WBC) count (**Figure 6**). Multiple lines of evidence support that the proteins encoded by these two genes interact to control immune response^87,88,89^, and some studies have promoted SLAM receptors as potential therapeutic targets for immune-mediated diseases^90^. Improved European genetic models of gene expression for whole blood also revealed an association between *PIGB* and heart failure, as well as *NOC3L* and asthma (**Figure 6**). Genetic variation in *PIGB* causes defects in glycosylphosphatidylinositol (GPI) biosynthesis^91^, which has been linked to cardiomyopathy from clinical outcome data^92^. The mammalian homolog of *NOC3L*, called *FAD24*, regulates the development of adipocytes^93^, which release adiponectin, a hormone that controls inflammation and is linked to asthma^94^.

Overall, our study has demonstrated several compelling applications and utilities of MAGEPRO. First, applying MAGEPRO gene expression prediction models to TWAS flags unstable disease/trait-associated genes by sometimes ablating significant associations generated by less accurate gene models. Second, MAGEPRO can help replicate European TWAS results in understudied ancestries, confirming population-shared gene-level effects on disease which has the potential to inform which European findings may be most clinically relevant to other populations. Third, utilizing MAGEPRO to perform TWAS in non-European populations can reveal population-specific gene-level disease effects. Fourth, MAGEPRO identifies biologically plausible novel connections between disease and putative gene-level risk factors, which previously could not be identified due to the lack of an available predictive *cis*-genetic gene model.

## Discussion

We developed a new method, MAGEPRO, that enhances population-specific gene expression prediction models by leveraging eQTL summary statistics from diverse ancestries and cell types. Briefly, MAGEPRO utilizes SuSiE to prioritize putative causal variants in external eQTL datasets, which are likely more informative than tagging variants when applied to the target population. We applied MAGEPRO to 8 eQTL cohorts representing 3 different ancestries, improving prediction accuracy by an average of 11% relative to LASSO and consistently outperforming all competing methods, including the state-of-the-art tool for genome-wide complex trait PRS using multi-ancestry data, PRS-CSx. The advantages offered by MAGEPRO were exemplied in small training cohorts (maximized improvement over conventional LASSO models), in low *cis*-heritable genes – which are more likely to be disease-critical, and in out-of-cohort prediction tasks for genetically similar populations.

When we applied MAGEPRO models to the TWAS framework, we identified 2,331 novel disease/trait-associated genes, including 1,350 as a result of improving (or adjusting) existing gene-trait associations and 981 that could not be identified by LASSO due to the lack of a predictive *cis*-genetic gene model. MAGEPRO identified several genes associated with white blood cell count that replicate across multiple ancestries, such as *PHTF1*, which is differentially expressed in leukemia patients. MAGEPRO also identified biologically plausible new associations, such as *PIGB* linked to heart failure, which has been evidenced by clinical outcome data.

We note several limitations to our work. First, MAGEPRO relies on the availability of target population genotype and gene expression data, which may be scarce for some ancestries (such as South Asians, South Americans, and others) and less accessible tissues. Second, MAGEPRO applies SuSiE to each external dataset independently, which may not be as powerful as modeling cross-ancestry or cross-tissue effect size correlations while fine-mapping. Third, MAGEPRO models are population-specific by design, which may complicate downstream analysis and limit generalizability when there are slight mismatches between the population structure of the training eQTL cohort and the target population (i.e., if the GWAS cohort has higher degrees of admixture). Fourth, while MAGEPRO definitively improves the accuracy of *cis*-genetic models of gene expression, limited availability of large ancestrally diverse GWAS continues to restrict the power of gene-disease association studies like TWAS. Despite these limitations, MAGEPRO is a powerful and robust method for creating population-specific *cis*-genetic models of gene expression and has provided clarifying and new insights related to the underlying risk factors of blood cell complex traits and immune-mediated diseases.

## Data Availability

This study did not generate any data that requires deposition in a repository. MAGEPRO software including documentation, tutorial, and code to reproduce analyses is publicly available at https://github.com/kaiakamatsu/MAGEPRO [DOI 10.5281/zenodo.13765893].

## Methods

### Baseline genetic model of gene expression: LASSO

We used the FUSION tool to build the standard gene expression prediction model, which uses individual-level genotype and gene expression data from a single target population (**Figure 1**, green box). In this baseline model, a single gene’s expression is modeled with standardized genotypes of *cis*-variants (within 1 Mb of the gene’s transcription start site (TSS)) in a multivariate linear regression:

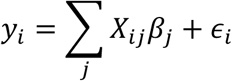

where for each individual *i*, *y_i_* is the gene expression of one gene, *j* indexes *cis*-variants, *X_ij_* is the standardized genotype of individual *i* at SNP *j*, *β_j_* is the true unobserved eQTL effect size, and *ε_i_* is the residual of gene expression not explained by modeled *cis*-genetic effects. We used LASSO (L1 norm) regularized linear regression from PLINK^95^ to estimate 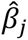 for each *cis*-variant such that we minimize the penalized sum of squares:

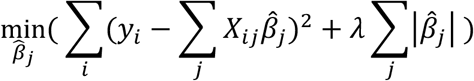

where *λ* is the sparsity parameter which is tuned via cross-validation. L1 regularization avoids overfitting by shrinking coefficients of less informative features (e.g., SNPs) to 0 and assigns nonzero coefficients to potentially predictive SNPs. When LASSO regression fails to find any meaningful predictors and pushes all coefficients to zero (potentially due to the limited sample size of the target population), we employ the “top 1” model as is done in the FUSION framework. The “top 1” model uses a single predictor SNP, specifically the SNP with the largest squared effect size from marginal *cis*-eQTL analysis. This approach systematically enables us to build a standard gene model for every gene in the analysis, to which we can compare MAGEPRO models informed by multiple ancestries.

### MAGEPRO (Multi-Ancestry Gene Expression Prediction Regularized Optimization)

MAGEPRO takes a three-step approach. First, it learns noisy estimates of SNP-gene effect sizes in the target population with a LASSO-regularized linear regression, identical to the baseline model described above (**Figure 1**, green box). Second, we apply the Sum of Single Effects (SuSiE) linear regression to each set of external eQTL summary statistics and we retain the posterior effect size estimates (**Figure 1**, blue box). SuSiE serves as a variable selection step, prioritizing potentially causal eQTLs which are more likely to be informative to the target population (see “Sum of Single Effects to prioritize variants from external summary statistics” section for more details regarding SuSiE). Finally, MAGEPRO models the gene expression of the target population as a function of the baseline LASSO-regularized model and the SuSiE posterior eQTL effect size estimates for each external dataset (**Figure 1**, white box):

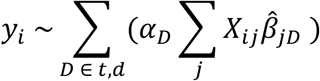

where for each individual *i*, *y_i_* is the gene expression of one gene, *D* indexes target (*t*) and external datasets (*d*), *j* indexes *cis*-variants, *X_ij_* is the standardized genotype of individual *i* at SNP *j*, 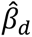 is a vector of posterior eQTL effect size estimates from external dataset *d*, and 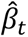 is a vector of estimated effect sizes from applying the baseline model described above to the target dataset. We used ridge (L2 norm) regression to fit 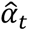 and 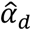; the dataset-specific mixing weights represent the relative contribution of each dataset to the prediction of gene expression, such that we minimize the loss function:

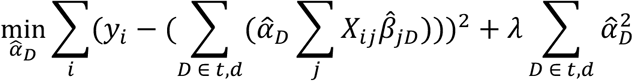

where *λ* is the sparsity parameter, which is tuned by ten-fold cross-validation^96^. We applied ridge regression to constrain the coefficients when two or more vectors are collinear, which may be common given that causal eQTL architecture is at least partially shared across populations.

### Simulations

We conducted simulations with various sample sizes and gene expression *cis*-heritability values to assess the robustness of MAGEPRO. We applied MAGEPRO, PRS-CSx, and LASSO to four predetermined levels of heritability (0.05, 0.1, 0.2, 0.4), which we confirmed using GCTA (**Supplementary Figure 22**). These heritability values were chosen based on the average estimated heritability values in quartiles of significantly heritable genes in LCL gene expression data from the GENOA African American (AA) population (0.088, 0.139, 0.202, 0.382). For each heritability value, we simulated 1,000 random genes and investigated the performance of each model across five target population (African) sample sizes (80, 160, 240, 400, 500). Simulated genotypes and gene expression levels for 500 EUR individuals (based on LD from the 1000 Genomes European ancestry group) and 500 AMR individuals (based on LD from the 1000 Genomes American ancestry group) were used to compute summary statistics, which we used as external datasets to apply MAGEPRO and PRS-CSx. Many of the functions that we used for our simulations are adopted from the Mancuso Lab TWAS simulator.

We assessed the performance of MAGEPRO in various simulated genetic architectures of gene expression: (1) the causal *cis*-eQTLs are the same across populations (same genomic position but not necessarily correlated in effect size), (2) the causal *cis*-eQTLs are different variants across populations but in high LD (*r*^2^ > 0.8), (3) true effect sizes of all shared causal *cis*-eQTLs are drawn independently across populations, and (4) true effect sizes of all shared causal *cis*-eQTLs are correlated across populations with effect size correlation set to 0.8, following recent work which estimated cis-molQTL (molecular quantitative trait loci) effect size correlations across ancestries^39^. The performances of LASSO, PRS-CSx, and MAGEPRO in simulations are evaluated with the prediction accuracy defined as 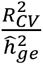. Please see the Supplementary Note section called “Simulation framework” for more details.

### Competing methods of gene expression prediction

We compare the performance of MAGEPRO against six different methods, capturing conventional methods applied to genome-wide complex trait data and gene expression data: meta-analysis, P+T, LASSO, SuSiE, Multipop, and PRS-CSx (see “*Baseline genetic model of gene expression: LASSO*” for more information on the LASSO model). We note that we do not compare the performance of elastic net or BLUP as recent work has shown that neither significantly outperform LASSO^39^.

The meta-analysis model refers to a sample-size weighted meta-analysis of all datasets, including the LASSO gene model which was developed using the training split of the target cohort. This strategy is commonly applied to GWAS data to maximize association power and identify shared effects.

P+T (pruning and thresholding) is an LD-informed pruning and p-value thresholding method^97^, also referred to as clumping and thresholding. Briefly, we iterate through SNPs in order of increasing p-value below a chosen threshold; p-values are computed from a marginal *cis*-eQTL analysis with the target cohort data. All variants in LD with the current SNP are removed until the iteration finishes. We performed a small grid-search across several LD *r*^2^ thresholds (0.2, 0.5, 0.8) and p-value thresholds (0.001, 0.01, 0.1, 0.5) to identify the pair of parameters that result in the best prediction result in 5-fold cross-validation. We performed P+T using PLINK and we used the target popuatlion genotypes as the in-sample LD reference panel.

SuSiE is the Sum of Single Effects regression model applied to the individual-level target population genotype and gene expression data. We used default parameters to run SuSiE (including a maximum number of allowed credible sets: L = 10, up to 100 iterative Bayesian stepwise selection (IBSS) iterations, and setting the estimated residual variance flag to TRUE if in-sample LD files were available and FALSE otherwise) and retained the resulting posterior effect size estimates to predict gene expression.

Multipop refers to a variation of MAGEPRO without the variable selection step using SuSiE. In this model, the raw external marginal *cis*-eQTL summary statistics are combined with the target population LASSO model using ridge regression. Benchmarking against this method allows us to evaluate if using SuSiE to prioritize potentially causal variants helps us create more accurate predictive models.

PRS-CSx is a Bayesian framework that improves cross-population polygenic prediction by learning an optimal linear combination of GWAS summary statistics from multiple ancestry groups to produce the final PRS. PRS-CSx employs a shared continuous shrinkage prior to SNP effects across populations (which assumes shared effects across populations) and leverages LD diversity across samples to enhance accuracy in effect size estimates. Although this method was originally designed to improve PRS for genome-wide complex traits and polygenic diseases in ancestrally diverse populations, we applied their command line tool to gene expression prediction to benchmark MAGEPRO. We utilized the shared shrinkage prior from PRS-CSx on the same datasets employed in MAGEPRO. Then, we learned an optimal linear combination of the post-shrinkage external datasets. To ensure that PRS-CSx utilizes the same features as MAGEPRO, we also added the LASSO gene model for the target population as one of the features in the linear combination. The authors of PRS-CSx recommend that the global shrinkage parameter, Φ, is adjusted based on the polygenicity of the phenotype. Since we expected the *cis*-genetic component of gene expression to be much less polygenic (involve fewer causal variants) than a genome-wide trait, we considered values of [10^-5^, 10^-6^, 10^-7^, 10^-8^, 10^-9^]. We applied PRS-CSx with these shrinkage parameters for 200 random genes with 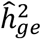 > 0 and 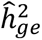 *p* < 0.05. We observed that gene model accuracy was robust across all values of Φ, and thus we selected the intermediate value (10^-7^) for the remaining analyses, which assumes that the polygenicity of *cis*-genetic gene expression regulation was well-represented by these 200 randomly selected genes (**Supplementary Figure 23**).

We note that BridgePRS^31^ is a recently published multi-ancestry PRS method that we considered for our study. However, their study demonstrated that BridgePRS only nominally outperforms PRS-CSx under highly polygenic genetic architectures, such as genome-wide complex traits. Therefore, we benchmarked MAGEPRO against PRS-CSx because we believed it was the best candidate among multi-ancestry PRS frameworks that are applicable to gene expression prediction.

### Preparing external summary statistics for MAGEPRO

We downloaded eQTL summary statistics from 5 publicly available datasets from 3 different ancestries including European, Latino/Hispanic and African American cohorts. For each dataset, we extracted full *cis*-eQTL summary statistics and filtered for 1,034,897 HapMap 3 SNPs included in GTEx. If the effect allele and alternate allele of the eQTLs were flipped in comparison to the target cohort SNPs, we multiplied the effect size of the eQTL from the external dataset by −1. We split each dataset into gene-specific files to facilitate downstream analysis with MAGEPRO. Dataset-specific preprocessing details are described in the Supplementary Note. To avoid overfitting, we utilized different combinations of external summary statistics depending on the target population to build the predictive model (**Table 1**).

### Sum of Single Effects model to prioritize variants from external summary statistics

We utilized the Sum of Single Effects regression model (SuSiE), specifically “SuSiE-RSS” (Regression with Summary Statistics), for variable selection from eQTL summary statistics data. SuSiE is a variable selection method that quantifies the uncertainty in which variables are selected by expressing the regression coefficients as a sum of single effects where only one of the variables has a nonzero coefficient. The model is fit with the IBSS procedure and produces posterior inclusion probabilities (PIPs) and posterior effect sizes for each SNP. The original SuSiE method requires individual-level phenotype and genotype data. In our MAGEPRO pipeline, external datasets only contain summary data, hence, we use SuSiE-RSS, which employs the “IBSS-ss” algorithm that relies only on sufficient statistics that can be approximated from the summary statistics. Within our pipeline, we conduct fine-mapping separately for each gene in each eQTL dataset. When available, we utilize in-sample correlation matrices (e.g., for MESA or GENOA datasets). In cases where in-sample matrices are not available, we employ out-of-cohort ancestry-matched alternatives (e.g., we used LD from the 1000 Genomes European population to fine-map the European eQTLGen dataset).

We note that the incorporation of the recently developed multi-ancestry statistical fine-mapping method, Sum of Shared Single Effects (SuShiE), may enhance the MAGEPRO framework by leveraging LD heterogeneity and modeling cross-ancestry effect size correlations to improve variable selection and effect size estimates in external eQTL datasets^39^. However, a version of SuShiE that is compatible with summary statistics was not released at the time of this study. Additionally, fine-mapping methods that are most compatible with MAGEPRO may also benefit from modeling cross-cell-type correlations to enable the sharing of information across eQTL datasets from different ancestries and cell types.

### Processing individual-level genotype and gene expression data

We used the same variant and relatedness filtering for all genotyping data, regardless of cohort. All genotype data processing was done using PLINK v1.9 and bcftools^98^. For the GENOA and MESA cohort, we imputed genotype data on the TOPMed server. Each ancestry/dataset assayed on different genotype platforms were imputed separately. The imputation was run using Minimac4 (1.8.0-beta4), using the TOPMed r3 reference panel and Eagle v2.4 phasing. We kept biallelic SNPs with high imputation quality (*r*^2^ > 0.9) for each imputed dataset and removed SNPs with MAF < 1%, Hardy Weinberg Equilbrium (HWE) *p* < 1 × 10^-6^, and genotyping rate < 1. We used plink (--rel-cutoff) to remove one individual of a pair that exhibited a relatedness greater than 0.05. When fitting the gene expression prediction models, we subset to HapMap 3 SNPs present in the dataset. Compared to keeping all SNPs in the genotype data, utilizing only HapMap 3 SNPs produces heritability estimates with smaller standard errors (**Supplementary Figure 24**).

The gene expression data for each cohort was inverse-normal transformed across individuals before fitting the gene expression prediction models^15^. We defined the *cis*-window of each gene as [start – 500 kilobases (Kb), end + 500 Kb]. The start and end positions were defined by gencode v26 gene annotations.

### Fitting gene expression prediction models

To calculate gene expression weights from real data, we used genotypes and gene expression data from whole blood and lung tissues of the GTEx cohort (EUR and AA populations), LCL gene expression data from GEUVADIS (EUR) and GENOA (AA), and monocyte gene expression data from MESA (EUR, AA, HIS) (**Table 1**). After extracting samples with both genotype and gene expression data, we performed imputation, variant-based filtering, and individual-level filtering steps described above. We regressed out the appropriate covariates from the gene expression data before fitting the gene expression prediction models. These covariates generally included 5 genotype PCs, genotype platform / site of data collection, sex, age, and gene expression PCs (depending on the sample size of the cohort). Please see the Supplementary Note for dataset-specific information.

The performance of gene expression prediction models in this paper are evaluated with 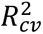 from a 5-fold cross validation. In each iteration of the cross-validation, we use the training split (4 folds) to learn a noisy estimate of *cis*-variant weights in a model identical to the standard gene expression prediction models described above. We include these weights from the training fold in a regularized linear combination with the other external datasets (consisting of SuSiE posterior effect sizes), and use the training split again to estimate the mixing weights 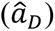. Finally, we extract the estimated coefficients and predict gene expression on the remaining testing split (5^th^ fold).

MAGEPRO computes both the target population SNP-gene weights 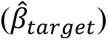 and the dataset mixture weights 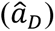 using the same training split. Therefore, we tested two potential training approaches: (1) the MAGEPRO training approach described above and (2) a training approach adopted from Márquez-Luna and colleagues^29^. In this second approach, we iteratively split the training samples (4 folds in 5-fold cross validation) into a 90% set used to estimate 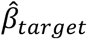 and computed the predicted gene expression for the 10% set (for each of the 10 folds). We then performed ridge regression across all training samples to estimate 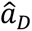 and finally re-estimated 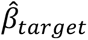 with the entire training split. We evaluated the two training approaches in predicting LCL gene expression at two different sample sizes (*n* = 100 and *n* = 346). We found that our MAGEPRO training approach outperformed the nested cross-validation approach in cross-validation prediction (*p* = 2 × 10^-90^ and *p* < 1 × 10^-200^ at *n* = 100 and *n* = 346, respectively, **Supplementary Figure 25**). While this could result from overfitting by MAGEPRO, we further compared the two approaches via an out-of-cohort prediction task in the GEUVADIS Yoruba (YRI) cohort. The gene models trained using the MAGEPRO approach exhibited higher accuracy (*p* = 0.01 and *p* = 4.9 × 10^-15^ at *n* = 100 and *n* = 346, respectively, **Supplementary Figure 25**). Therefore, we concluded that our training approach that utilizes the same training split to estimate both 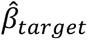 and 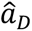 is valid.

### Validation of MAGEPRO models out-of-cohort

We validate the improved MAGEPRO models by training our models in one cohort and applying them to a different cohort of a similar ancestry and cell type. To facilitate the application of gene expression prediction models across datasets, we subset to SNPs in common between the two datasets within each ancestry. Without this additional SNP-based filtering step, we risk creating predictive models that assign a non-zero effect size to SNPs that are not present in the out-of-cohort validation set.

To validate the LCL gene models in the European population, we built predictive models in the GEUVADIS population and validated them in the GENOA population. We worked with 718,414 HapMap 3 SNPs that are present among GEUVADIS European individuals and GENOA European American individuals.

For individuals of African American descent, we built predictive models in the GENOA population and validated them in the GEUVADIS YRI (Yoruba) population. We worked with 718,838 HapMap 3 SNPs that are present among GENOA African American individuals and GEUVADIS YRI individuals.

### TWAS using GWAS summary statistics

We collected GWAS summary statistics for 15 blood cell traits from a previous study^60^ (AFR *N* = 13,391, EUR *N* = 516,979, HIS *N* = 6,849) and 7 immune-mediated diseases from the Global Biobank Meta-analysis Initiative (GBMI) (AFR *N* = 26,052, EUR *N* = 1,024,298, AMR *N* = 15,490). We updated the variant identifiers to dbSNP v151 and used the munge_sumstats.py script from LD score regression^99^ to perform quality control and filtering. We evaluated the TWAS results for the union of significantly heritable genes across populations (LCL: 6,872 genes, Monocyte: 5,920 genes, Lung: 8,807 genes) that have gene models that explain some proportion of variance in gene expression (*R*^2^ > 0, *p* < 0.05). TWAS p-values were subjected to a Bonferroni significance threshold to account for multiple hypothesis testing.

### Statistics and Reproducibility

First, as described above, we filtered each external eQTL dataset and target cohort genotypes to HapMap 3 SNPs. Second, we evaluated the performance of MAGEPRO on significantly heritable genes (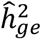 > 0, *p* < 0.01) with eQTL data from at least 1 external dataset. Third, as described above, we performed random down-sampling of certain cohorts to test MAGEPRO at smaller sample sizes. Fourth, as described above, we evaluated TWAS results from gene models that explain some proportion of variance in gene expression (*R*^2^ > 0, *p* < 0.05) to prevent spurious associations from estimated eQTL effect sizes that poorly capture gene expression regulation. Fifth, as described above, 1,000 random genes were simulated for each genetic architecture to robustly evaluate MAGEPRO performance. Randomization and blinding were not pertinent to our study.

## Data Availability

Blood trait GWAS summary statistics are available at http://www.mhi-humangenetics.org/en/resources/. Immune-related disease GWAS summary statistics are available at https://www.globalbiobankmeta.org/resources. GTEx gene expression and genotype data were acquired from dbGaP accession phs000424.v9.p2. MESA genotype data was acquired from dbGaP accession phs000209.v13.p3 (file names: phg000071.v2.NHLBI_SHARE_MESA.genotype-calls-matrixfmt.c1 and phg000071.v2.NHLBI_SHARE_MESA.genotype-calls-matrixfmt.c2), GENOA genotype data was acquired from dbGaP accession phs001238.v2.p1, and GEUVADIS genotype data is publicaly available at https://www.ebi.ac.uk/biostudies/arrayexpress/studies/E-GEUV-1. MESA gene expression data was acquired from NCBI GEO accession GSE56045, GENOA gene expression data was acquired from NCBI GEO accessions GSE138914 (African American individuals) and GSE49531 (European individuals), and GEUVADIS gene expression data is publicly available at https://uchicago.app.box.com/s/ewnrqs31ivobz2sn6462cq2eb423dvpr. 1000 Genomes LD reference files were acquired from https://www.bridgeprs.net/guide_input/.

As described in https://github.com/kaiakamatsu/MAGEPRO/tree/main/PROCESS_DATASET, all eQTL summary statistics were publicly available: eQTLGen (https://molgenis26.gcc.rug.nl/downloads/eqtlgen/cis-eqtl/SMR_formatted/cis-eQTL-SMR_20191212.tar.gz), GTEx (https://console.cloud.google.com/storage/browser/gtex-resources;tab=objects?prefix=&forceOnObjectsSortingFiltering=false), GENOA (http://www.xzlab.org/data/AA_summary_statistics.txt.gz), and MESA (https://www.dropbox.com/sh/f6un5evevyvvyl9/AAA3sfa1DgqY67tx4q36P341a?dl=0).

## Code Availability

MAGEPRO software including documentation and tutorial is publicly available at https://github.com/kaiakamatsu/MAGEPRO [DOI <u>10.5281/zenodo.13765893</u>]. The Mancuso Lab TWAS Simulator is available at https://github.com/mancusolab/twas_sim. The FUSION software is available at http://gusevlab.org/projects/fusion. PRS-CSx is available at https://github.com/getian107/PRScsx. SuSiE is available as an R package and it is described at https://stephenslab.github.io/susieR/index.html. The munge_sumstats.py script is available in the LDSC github at https://github.com/bulik/ldsc/tree/master. To improve the runtime of MAGEPRO, we utilized GNU Parallel available at https://zenodo.org/records/10901541.

## Acknowledgements

This work was supported by funding from the National Science Foundation (NSF) (Award #2336469 awarded to T.A.) and the National Institutes of Health (NIH) (NHGRI R01HG013671 awarded to T.A.). The funders played no role in study design, data collection and analysis, decision to publish or preparation of the manuscript. Support for GENOA was provided by the National Heart, Lung and Blood Institute (HL054457, HL054464, HL054481, HL119443, and HL087660) of the National Institutes of Health. We would like the thank the Mayo Clinic Genotyping Core, the DNA Sequencing and Gene Analysis Center at the University of Washington, and the Broad Institute for their genotyping and sequencing services. We would also like to thank the GENOA participants. This manuscript was not prepared in collaboration with investigators from the Genetic Epidemiology Network of Arteriopathy and does not necessarily reflect the opinions or views of the Genetic Epidemiology Network of Arteriopathy or NHLBI. MESA and the MESA SHARe project are conducted and supported by the National Heart, Lung, and Blood Institute (NHLBI) in collaboration with MESA investigators. Support for MESA is provided by contracts N01-HC95159, N01-HC-95160, N01-HC-95161, N01-HC-95162, N01-HC-95163, N01-HC-95164, N01-HC-95165, N01-HC95166, N01-HC-95167, N01-HC-95168, N01-HC-95169 and CTSA UL1-RR-024156. The Genotype-Tissue Expression (GTEx) Project was supported by the Common Fund of the Office of the Director of the National Institutes of Health, and by NCI, NHGRI, NHLBI, NIDA, NIMH, and NINDS. This work used the Expanse HPC server at the San Diego Supercomputer Center (SDSC) through allocation BIO230210 from the Advanced Cyberinfrastructure Coordination Ecosystem: Services & Support (ACCESS) program, which is supported by National Science Foundation grants #2138259, #2138286, #2138307, #2137603, and #2138296.

## Author Contributions

K.A. and T.A. conceived and designed the study. K.A. conducted simulation analyses. K.A. and and S.G. conducted real data analysis. T.A. managed GTEx, GENOA, and MESA data through dbGaP. K.A., S.G., and T.A. wrote the initial draft of the manuscript and contributed to the final manuscript.

