## Supplementary Note for "Powerful mapping of *cis*-genetic effects on gene expression across diverse populations reveals novel disease-critical genes"

### Supplementary Table Captions

**Supplementary Table 1. Numerical results for the average accuracy of gene expression prediction models in simulations.** We evaluate the gene expression prediction accuracy ($\frac{R_{cv}^2}{\hat{h}_{ge}^2}$) of LASSO, PRS-CSx, and MAGEPRO across five different sample sizes and four different preset heritability values in simulations. We performed simulations with four shared causal variants with correlated effect sizes across populations (r = 0.8). Accuracy is calculated as the ratio of the cross-validation $R_{cv}^2$ and the GCTA-estimated *cis*-heritability of gene expression ($\hat{h}_{ge}^2$). The standard errors of the mean are computed as the standard deviation of the accuracy values across simulations, divided by the square root of the number of simulations (e.g., 1,000).

**Supplementary Table 2. Test statistics from comparing the accuracy of MAGEPRO models to LASSO and PRS-CSx in simulations.** We compare the gene expression prediction accuracy ($\frac{R_{cv}^2}{\hat{h}_{ge}^2}$) of MAGEPRO to LASSO and PRS-CSx across five different sample sizes and four different preset heritability values in simulations. We performed simulations with four shared causal variants with correlated effect sizes across populations (r = 0.8). We report the t-statistic, p-value, and difference of means from a two-sided paired t-test.

**Supplementary Table 3. Numerical results for the average squared error of effect sizes of causal variants in simulations.** We evaluate the squared error of causal variant effect sizes for LASSO, PRS-CSx, and MAGEPRO models across five sample sizes and preset heritability of 10% in simulations. We performed simulations with four shared causal variants with correlated effect sizes across populations (r = 0.8). The squared error is calculated as the squared difference between the actual effect size and estimated effect size. The standard errors of the mean are computed as the standard deviation of measurements across simulations, divided by the square root of the number of simulations (e.g., 1,000).

**Supplementary Table 4. Test statistics from comparing the squared error of effect sizes of causal variants in simulations.** We compared the squared error of causal variant effect sizes for LASSO, PRS-CSx, and MAGEPRO models across five sample sizes and preset heritability of 10% in simulations. We performed simulations with four shared causal variants with correlated effect sizes across populations (r = 0.8). The squared error is calculated as the squared difference between the actual effect size and estimated effect size. We report the t-statistic, p-value, and difference of means from a two-sided paired t-test.

**Supplementary Table 5. Numerical results for the average accuracy of gene expression prediction models while increasing the number of causal variants in simulations.** We evaluate the gene expression prediction accuracy ($\frac{R_{cv}^2}{\hat{h}_{ge}^2}$) of LASSO, PRS-CSx, and MAGEPRO models across varying numbers of causal variants, while keeping the sample size fixed at 240 and heritability at 10%. We performed simulations with four shared causal variants with correlated effect sizes across populations (r = 0.8). Accuracy is calculated as the ratio of the cross-validation $R_{cv}^2$ and the GCTA-estimated *cis*-heritability of gene expression ($\hat{h}_{ge}^2$). The standard errors of the mean are computed as the standard deviation of measurements across simulations, divided by the square root of the number of simulations (e.g., 1,000).

**Supplementary Table 6. Test statistics from comparing the accuracy of gene expression prediction models while increasing the number of causal variants in simulations.** We compare the gene expression prediction accuracy ($\frac{R_{cv}^2}{\hat{h}_{ge}^2}$) of LASSO, PRS-CSx, and MAGEPRO models across varying numbers of causal variants, while keeping the sample size fixed at 240 and heritability at 10%. We performed simulations with four shared causal variants with correlated effect sizes across populations (r = 0.8). We report the t-statistic, p-value, and difference of means from a two-sided paired t-test.

**Supplementary Table 7. Numerical results for the average accuracy of gene expression prediction models in real data.** We evaluate the accuracy of seven gene expression prediction methods in predicting LCL gene expression in the GENOA AA cohort ($n = 346$). We computed an average across 4,141 *cis*-heritable genes. The standard errors of the mean are computed as the standard deviation of accuracy across genes, divided by the square root of the number of genes (e.g., 4,141).

**Supplementary Table 8. Numerical results for the average accuracy of gene expression prediction models in real data with 100 training samples.** We evaluate the accuracy of seven gene expression prediction methods in predicting LCL gene expression in 100 randomly sampled individuals from the GENOA AA cohort. We computed an average across 977 *cis*-heritable genes. The standard errors of the mean are computed as the standard deviation of accuracy across genes, divided by the square root of the number of genes (e.g., 977).

**Supplementary Table 9. Performance of gene expression prediction models relative to LASSO across 10 target datasets.** We report the percent change in gene expression prediction accuracy ($\frac{R_{cv}^2}{\hat{h}_{ge}^2}$) of the top five methods relative to LASSO across ten different eQTL cohorts. Accuracy is calculated as the ratio of the cross-validation $R_{cv}^2$ to the GCTA-estimated *cis*-heritability of gene expression ($\hat{h}_{ge}^2$).

**Supplementary Table 10. Numerical results for the average change in accuracy relative to LASSO across genes grouped by GCTA-estimated *cis*-heritability.** We report the average change in accuracy between MAGEPRO and LASSO (MAGEPRO $\frac{R_{cv}^2}{\hat{h}_{ge}^2}$ - LASSO $\frac{R_{cv}^2}{\hat{h}_{ge}^2}$) across 10 quantiles of genes grouped by GCTA-estimated *cis*-heritability. Accuracy is calculated as the ratio of the cross-validation $R_{cv}^2$ to the GCTA-estimated *cis*-heritability of gene expression ($\hat{h}_{ge}^2$). The standard errors of the mean are computed as the standard deviation of the change in accuracy across genes, divided by the square root of the number of genes.

**Supplementary Table 11. Phenotypes analyzed in TWAS.** This table presents 22 diseases/traits selected for TWAS analysis, including 15 blood traits from Chen et al. 2020 *Cell* and 7 immune-mediated diseases from the Global Biobank Meta-analysis Initiative (GBMI). We conducted TWAS using GWAS summary statistics across three distinct ancestries. We report the trait abbreviation and name.

**Supplementary Table 12. eQTL effect sizes and GWAS z-scores of *ZNF213-AS1*.** We report the eQTL effect sizes of the MAGEPRO, LASSO, and SuSiE models and the GWAS z-score of each SNP. Some GWAS z-scores are imputed, identical to the FUSION TWAS framework. The data does not encompass the entire *cis*-window of the gene because certain regions lack eQTL signals.

**Supplementary Table 13. eQTL effect sizes and GWAS z-scores of *RGS14*.** We report the eQTL effect sizes of the MAGEPRO and LASSO models and the GWAS z score of each SNP. Some GWAS z scores are imputed, identical to the FUSION TWAS framework. The data does not encompass the entire *cis*-window of the gene because certain regions lack eQTL signals.

**Supplementary Table 14. New gene-trait associations from MAGEPRO not previously found by LASSO.** We report significant gene-trait associations uniquely identified by MAGEPRO, but not by LASSO, across 66 GWAS summary statistics and 7 eQTL datasets in the TWAS analysis. Only genes with a gene expression prediction $R_{cv}^2 > 0$ in all methods are analyzed. We determined the Bonferroni significance threshold separately for each eQTL dataset ($p < \frac{0.05}{\#\ genes\ tested\ in\ dataset}$).

**Supplementary Table 15. New gene-trait associations from MAGEPRO not previously found by any other model.** We report significant gene-trait associations identified exclusively by MAGEPRO, but not by any other models (LASSO, SuSiE, PRS-CSx), across 66 GWAS summary statistics and 7 eQTL datasets in the TWAS analysis. Only genes with a gene expression prediction $R_{cv}^2$ significantly greater than 0 in all methods are analyzed. We determined the Bonferroni significance threshold separately for each eQTL dataset ($p < \frac{0.05}{\#\ genes\ tested\ in\ dataset}$).

**Supplementary Table 16. Results from applying Monocyte African American MAGEPRO and LASSO models to TWAS in 15 blood cell traits.** We present TWAS results from MAGEPRO and LASSO for 15 blood cell traits, using genetic models of gene expression trained on Monocyte gene expression data from the MESA African American cohort. We analyzed genes that have a cross-validation $R^2_{cv}$ significantly greater than 0 in both MAGEPRO and LASSO. The nominal significance threshold is $p < 0.05$ and Bonferroni significance threshold is $p < \frac{0.05}{5920}$.

**Supplementary Table 17. Results from applying Monocyte African American MAGEPRO and PRS-CSx models to TWAS in 15 blood cell traits.** We present TWAS results from MAGEPRO and PRS-CSx for 15 blood cell traits, using genetic models of gene expression trained on Monocyte gene expression data from the GENOA African American cohort. We analyzed genes that have a cross-validation $R^2_{cv}$ significantly greater than 0 in both MAGEPRO and PRS-CSx. The nominal significance threshold is $p < 0.05$ and Bonferroni significance threshold is $p < \frac{0.05}{5920}$.

**Supplementary Table 18. Genes associated with white blood cell count across multiple populations.** We report TWAS results from MAGEPRO models for genes that are significantly associated with white blood cell count across at least two ancestries. Significance is determined with a Bonferroni threshold of $p < \frac{0.05}{5920}$.

**Supplementary Table 19. TWAS associations found specifically in non-European populations.** We report significant gene-trait associations uniquely identified in non-European TWAS, but not detected in European TWAS, across 66 GWAS summary statistics and 7 eQTL datasets. Genes with a gene expression prediction $R^2_{cv}$ significantly greater than 0 in both LASSO and MAGEPRO were analyzed, and the better-performing model was selected for TWAS. We determined the Bonferroni significance threshold separately for each eQTL dataset ($p < \frac{0.05}{\#\ genes\ tested\ in\ dataset}$).

**Supplementary Table 20. TWAS associations from new gene models created by MAGEPRO.** We report significant gene-trait associations identified by MAGEPRO models for genes that lacked a predictive LASSO model ($R^2_{cv}$ not significantly positive), aggregated across TWAS using 66 GWAS summary statistics and 7 eQTL datasets. We determined the Bonferroni significance threshold separately for each eQTL dataset ($p < \frac{0.05}{\#\ genes\ tested\ in\ dataset}$). Columns with NA values for TWAS statistics resulting from the LASSO model indicate that the gene was not included in the TWAS analysis because its gene model did not show a significantly positive $R^2$.

### Supplementary Note

#### *Simulation framework*

#### A. Simulating Genotypes

To simulate the genotypes, we used 1000 Genomes (1KG) Phase 3 genotypes to represent the LD structure of each population. To construct a simulated gene, we chose a random variant from the EUR 1KG reference files on a randomly selected chromosome and extracted variants in a 1Mb region centered around the chosen SNP. We filtered the variants for HapMap 3 SNPs present in GTEx to ensure that our simulations are similar to the application of MAGEPRO to real data, where HapMap 3 SNPs present in GTEx is a standard that we applied to all analyzed datasets. To ensure an adequate number of variants in the *cis*-region, we confirmed that the simulated gene has at least 50 variants. We simulated individual-level genotypes of EUR, AMR and AFR individuals using the 1KG samples as an LD reference panel. The genotypes were randomly drawn from a standard normal distribution and integrated with the LD matrix.

#### B. Simulating Gene Expression

For each population, we simulated gene expression as a function of the *cis*-genetic component of heritability and non-genetic noise:

$$y = X\beta + \varepsilon$$

where X is a simulated standardized genotype matrix ($n \times p$) of $n$ people and $p$ variants, $\beta$ is a length $p$ vector of effect sizes in which a randomly selected causal variant has a nonzero effect size drawn from a normal distribution with mean 0 and variance $h_{ge}^2$ (*cis*-genetic heritability of gene expression), and $\varepsilon$ is the non-genetic noise drawn from a distribution with mean 0 and variance $1 - h_{ge}^2$. We set $h_{ge}^2$ to four different values (0.05, 0.1, 0.2, 0.4) for our simulated genes and estimated the heritability with GCTA to confirm that it corresponds to the value we assigned. To test the performance of MAGEPRO in various genetic architectures, we also simulated genes with multiple causal eQTLs. To this end, we randomly selected $c$ SNPs to have a nonzero effect size drawn from a normal distribution with mean 0 and variance $\frac{h_{ge}^2}{c}$.

#### C. Simulating correlated causal eQTL effect sizes across ancestries

There are lines of evidence to show that effect sizes of causal molecular QTLs (molQTLs) are correlated across ancestries. Therefore, we also tested the performance of each gene expression prediction model while controlling the correlation of causal effect sizes across ancestries. First, we define two random effect sizes from a centered normal distribution with variance $h_{ge}^2$.

$$\beta_{population\ 1} \sim N\left(0, \sqrt{h_{ge}^2}\right)$$

$$\beta_{temporary} \sim N\left(0, \sqrt{h_{ge}^2}\right)$$

$\beta_{population\ 1}$ is the causal eQTL effect size for one population, while $\beta_{temporary}$ is a value that is sampled independently from $\beta_{population\ 1}$. Our goal is to compute $\beta_{population\ 2}$ with variance $h_{ge}^2$ and also a correlation with $\beta_{population\ 1}$ of $r$.

$$\beta_{population\ 2} \sim N(0, \sqrt{h_{ge}^2})$$

$$Corr(\beta_{population\ 1}, \beta_{population\ 2}) = r$$

To this end, we define $\beta_{population\ 2}$ as the following:

$$\beta_{population\ 2} = r\beta_{population\ 1} + \sqrt{1-r^2}\beta_{temporary}$$

We confirm the (1) mean of $\beta_{population\ 2}$, (2) variance of $\beta_{population\ 2}$, and (3) correlation between $\beta_{population\ 1}$ and $\beta_{population\ 2}$.

(1) The mean of $\beta_{population\ 2}$ is 0 due to the linearity of expectation.

$$E[\beta_{population\ 2}] = E\left[r\beta_{population\ 1} + \sqrt{1-r^2}\beta_{temporary}\right]$$
$$= rE[\beta_{population\ 1}] + \sqrt{1-r^2}E[\beta_{temporary}]$$
$$= r(0) + \sqrt{1-r^2}(0) = 0$$

(2) The variance of $\beta_{population\ 2}$is also simple because $\beta_{population\ 1}$ and $\beta_{temporary}$ are independent.

$$Var(\beta_{population\ 2}) = Var\left(r\beta_{population\ 1} + \sqrt{1-r^2}\beta_{temporary}\right)$$
$$= r^2Var(\beta_{population\ 1}) + (1-r^2)Var(\beta_{temporary})$$
$$= r^2h_{ge}^2 + (1-r^2)h_{ge}^2$$
$$= h_{ge}^2(r^2 + 1 - r^2) = h_{ge}^2$$

(3) The correlation between $\beta_{population\ 1}$ and $\beta_{population\ 2}$ can be confirmed using some properties of covariance.

$$Cov(\beta_{population\ 1}, \beta_{population\ 2}) = Cov\left(\beta_{population\ 1}, r\beta_{population\ 1} + \sqrt{1-r^2}\beta_{temporary}\right)$$
$$= Cov(\beta_{population\ 1}, r\beta_{population\ 1}) + Cov\left(\beta_{population\ 1}, \sqrt{1-r^2}\beta_{temporary}\right)$$
$$= r\,Cov(\beta_{population\ 1}, \beta_{population\ 1}) + \sqrt{1-r^2}Cov(\beta_{population\ 1}, \beta_{temporary})$$
$$= rVar(\beta_{population\ 1}) + \sqrt{1-r^2}(0) = rh_{ge}^2$$
$$Corr(\beta_{population\ 1}, \beta_{population\ 2}) = \frac{Cov(\beta_{population\ 1}, \beta_{population\ 2})}{\sigma_{\beta_{population\ 1}}\sigma_{\beta_{population\ 2}}}$$
$$= \frac{rh_{ge}^2}{\sqrt{h_{ge}^2}\sqrt{h_{ge}^2}} = r$$

In our simulation analysis, we set the correlation of causal eQTL effect sizes across ancestries to 0.8.

**D. eQTL Summary Statistics and SuSiE in simulations**

We used the simulated genotypes and gene expression of the EUR and AMR 1KG cohorts to simulate eQTL summary statistics as external datasets used to inform prediction in the target simulated African 1KG population. Standardized gene expression values across individuals were regressed on the genotypes of each variant individually in a linear regression, and the coefficient (estimated effect size) and p-value were extracted. Signed SNP-SNP correlation matrices created from simulated genotypes (representing LD structure) and these simulated eQTL summary statistics were used as inputs to SuSiE. The posterior effect size estimates from SuSiE were used as features in our MAGEPRO model.

***Dataset-specific processing of individual-level genotype and gene expression data for fitting genetic models of gene expression***

GTEx: we downloaded genotype and whole blood gene expression data of 80 African American individuals and 568 European individuals from GTEx (dbGAP accession phs000424.v9.p2). The dataset included gene expression data for 19,602 genes expressed in the whole blood tissue. 5 genotype principal components (PCs), PCR platform (T/F), sex, and several PEER factors were regressed out of the gene expression levels before the analysis. We adjusted the number of PEER factors based on the sample size of the target population (60 PEER factors for EUR, 15 PEER factors for AA) (https://github.com/broadinstitute/gtex-pipeline/tree/master/qtl).

GEUVADIS: we downloaded publicly available genotype and LCL gene expression data of 373 European individuals and 89 Yoruba (YRI) individuals from the 1000 Genomes Project. After downloading the genotype data, we performed the same variant-based and individual-level filtering steps described in the main text. This resulted in 1,006,722 SNPs for 364 European individuals and 904,740 SNPs for 89 Yoruba individuals. We regressed out the following covariates from the gene expression data: sex, 5 genotyping PCs, and gene expression PCs (30 for EUR, 15 for YRI).

GENOA: we downloaded genotype data for 1,532 African American individuals and 1,507 European American individuals (dbGaP accession phs001238.v2.p1). We downloaded LCL gene expression data from NCBI GEO (GSE49531 for EUR, GSE138914 for AA). We took the intersection of individuals in the genotype data and the gene expression data. After downloading the data, we performed the same imputation, variant-based filtering, and individual-level filtering steps described in the main text. This resulted in 728,670 SNPs for 279 European American individuals and 810,927 SNPs for 346 African American individuals. We regressed out the following covariates from the gene expression data: genotyping platform, age, sex, 5 genotyping PCs, and 30 gene expression PCs.

MESA: we downloaded genotype and monocyte gene expression data for 582 Caucasian (EUR) individuals, 386 Hispanic (HIS) individuals, and 234 African American (AA) individuals from dbGaP accession phs000209.v13.p3 and NCBI GEO GSE56045. We performed the same imputation, variant-based filtering, and individual-level filtering steps described in the main text. This resulted in 597,137 SNPs for 574 EUR individuals, 601,862 SNPs for 242 HIS individuals, and 565,338 SNPs for 224 AA individuals. For the gene expression data, Illumina IDs were converted to Ensembl IDs. If multiple Illumina IDs corresponded to the same Ensembl ID, we used the average of the expression values. We regressed out the following covariates from the gene expression data: age, sex, site of data collection, cell type contamination (B cells, T cells, NK cell, Neutrophils), 5 genotyping PCs, and 30 gene expression PCs.

#### ***Computational efficiency of MAGEPRO***

It is often desirable to compute genetic models of gene expression across thousands of genes for a single analysis. Therefore, we implemented MAGEPRO to be able to perform computations on batches of genes in parallel across multiple HPC cluster jobs (--batch and --num_batches). Additionally, our tool allows users to specify the number of threads to use within each batch, where each thread can process one gene at a time (--n_threads). Adjusting the number of threads to utilize will accelerate the runtime on both HPC clusters (multiple threads per batched job) and local computer (multiple threads on the user's device). For example, allocating 4 threads results in a 3.4x speed up (1549 seconds to 452 seconds) in processing 100 randomly selected genes (**Supplementary Figure 17**).

#### ***SuSiE diagnostics and recommendations***

We encountered some unexpected results when testing SuSiE-RSS (Regression using Summary Statistics) in our method, such as large posterior effect size estimates ($\sum \hat{\beta}_j^{\,2} > 1$) and difficulty converging in the default number of IBSS iterations (e.g., 100). Our MAGEPRO pipeline is designed to create diagnostic plots when these potential issues are encountered. We suggest users consult this diagnostic plot and identify potential sources of error, such as mismatch between the LD reference file and target population.

### Supplementary Figures

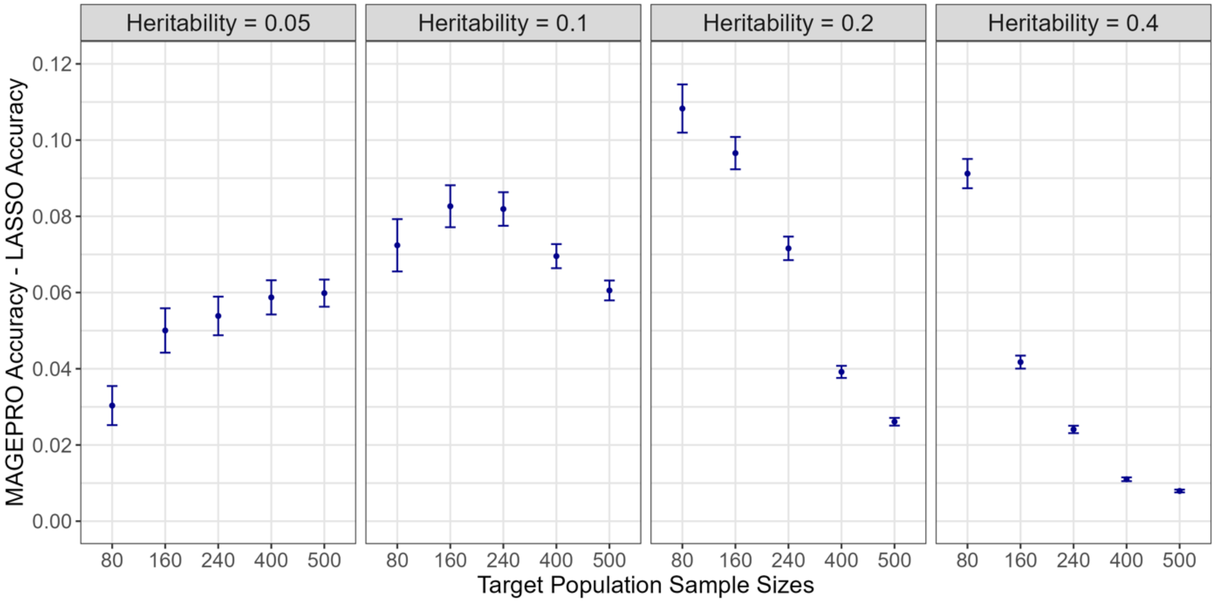


**Supplementary Figure 1. Improvement in prediction accuracy provided by MAGEPRO relative to LASSO.** Data plotted are the mean differences in prediction accuracy $(\frac{R^2_{CV}}{\hat{h}^2_{ge}})$ between MAGEPRO and LASSO across 1,000 independently simulated genes, where confidence intervals represent $\pm$ 1 standard error. Simulations were run with 4 causal variants shared across ancestries with correlated effect sizes (r = 0.8).

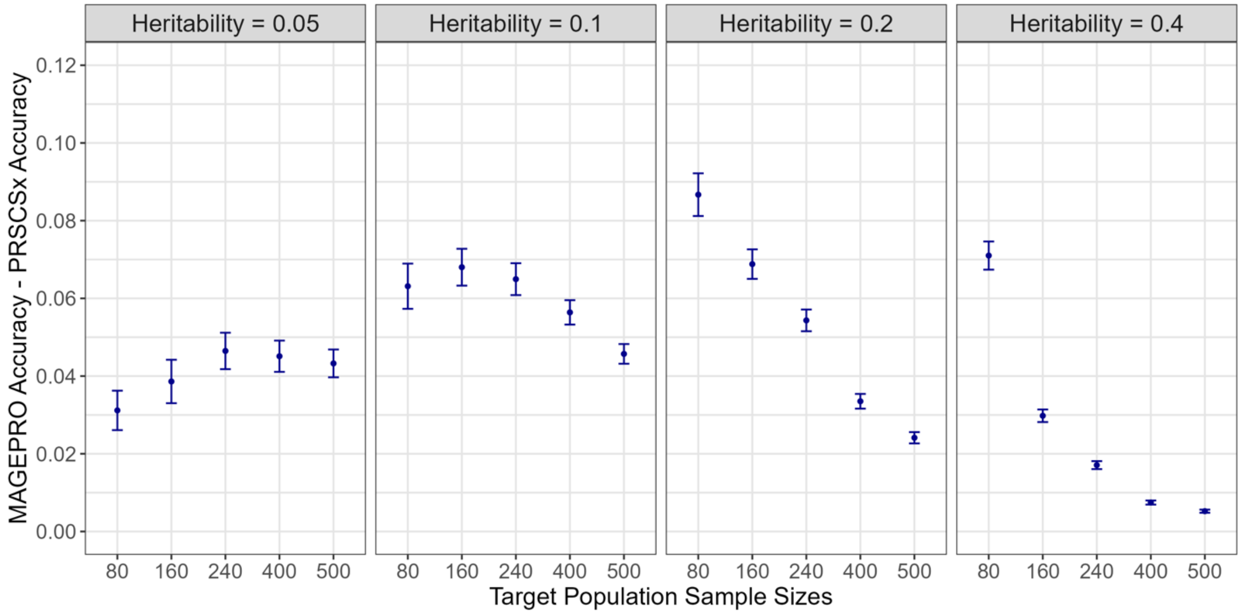


**Supplementary Figure 2. Improvement in prediction accuracy provided by MAGEPRO relative to PRS-CSx.** Data plotted are the mean differences in prediction accuracy ($\frac{R_{CV}^2}{\hat{h}_{ge}^2}$) between MAGEPRO and PRS-CSx across 1,000 independently simulated genes, where confidence intervals represent $\pm$ 1 standard error. Simulations were run with 4 causal variants shared across ancestries with correlated effect sizes (r = 0.8).

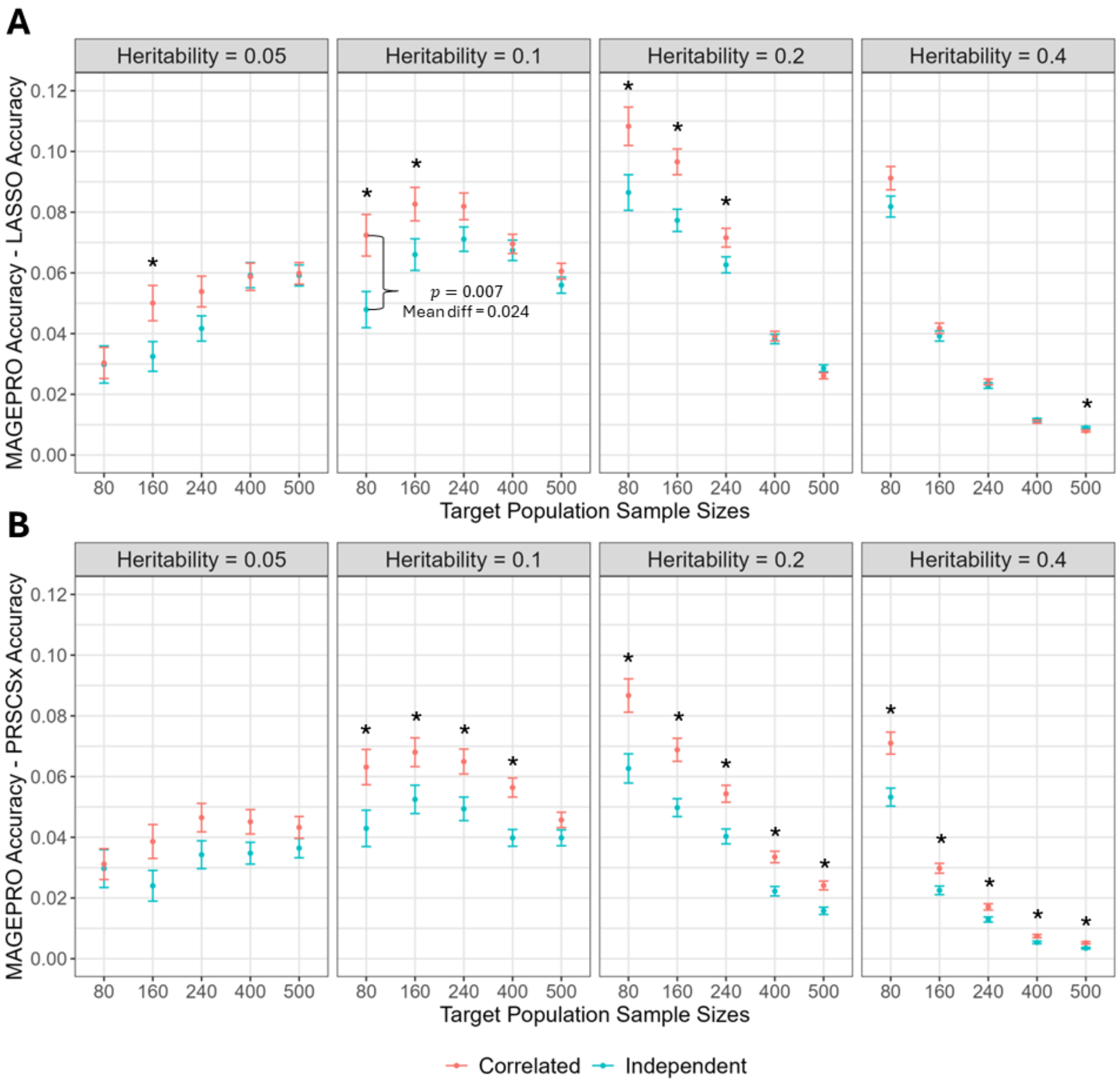


**Supplementary Figure 3. Comparison of MAGEPRO to LASSO and PRS-CSx when causal variant effect sizes across ancestries are correlated (r = 0.8) and when they are independent.**

Data plotted are the mean differences in prediction accuracy ($\frac{R^2_{CV}}{\hat{h}^2_{ge}}$) between MAGEPRO and (A) LASSO or (B) PRS-CSx across 1,000 independently simulated genes, where confidence intervals represent $\pm$ 1 standard error. In each simulation, all 4 causal variants were shared across populations. An asterisk (*) above a pair of results indicates a significant difference of means ($p < 0.05$ from a two-sided unpaired t-test).

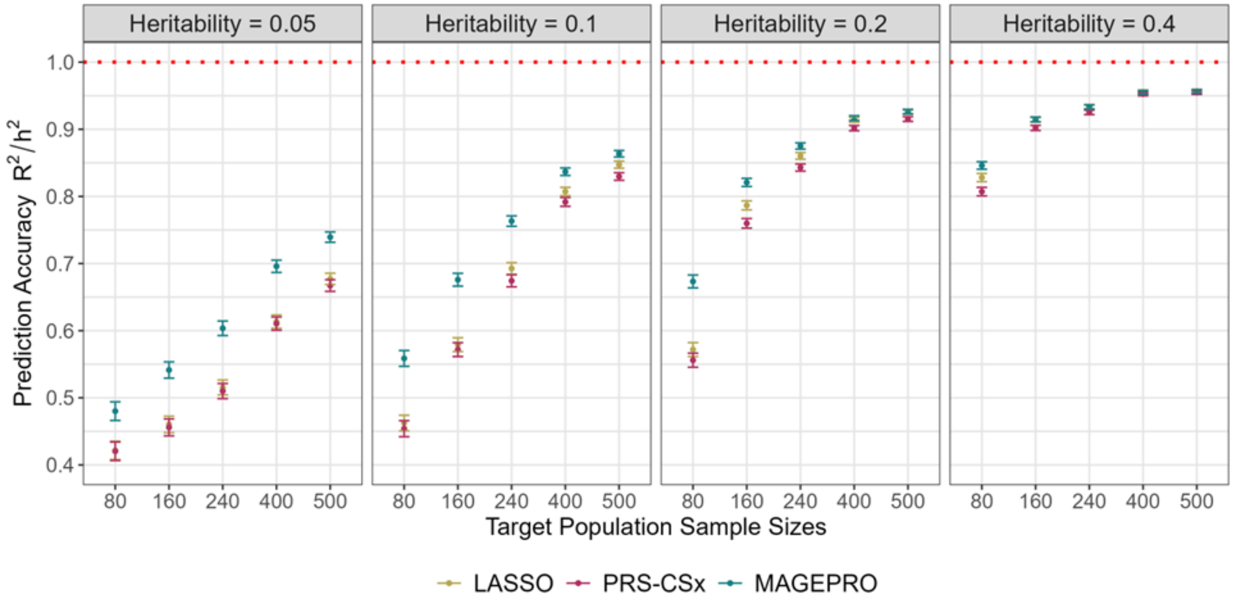


**Supplementary Figure 4. Performance of gene expression prediction models when the causal variant is different across populations but in high LD relative to the target cohort.** Gene expression was simulated from a single causal variant. Data plotted are the mean prediction accuracy ($\frac{R_{CV}^2}{\hat{h}_{ge}^2}$) across 1,000 independently simulated genes, where confidence intervals represent $\pm$ 1 standard error.

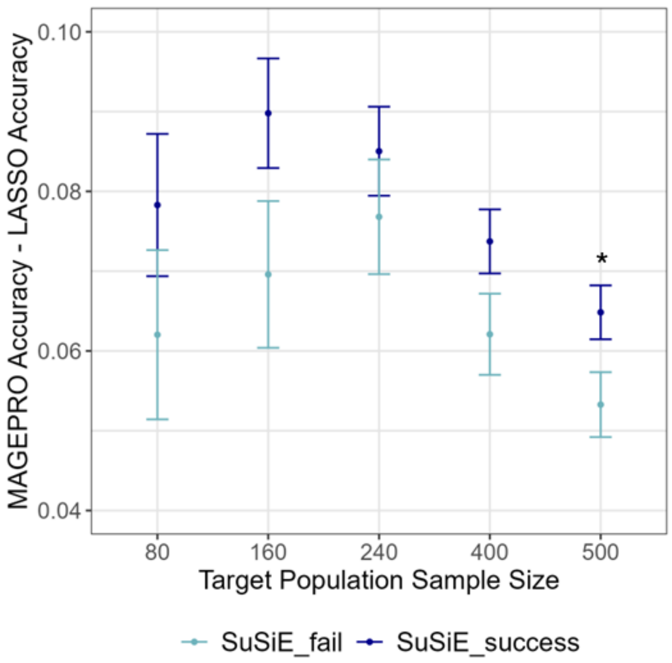


**Supplementary Figure 5. Performance of MAGEPRO model when SuSiE successfully identifies at least 1 causal variant from the external datasets ($PIP \geq 0.95$).** "SuSiE_success" refers to the scenario when SuSiE assigns at least 1 variant a $PIP \geq 0.95$ in either of the two external datasets. Data plotted are the mean differences in prediction accuracy ($\frac{R^2_{CV}}{\hat{h}^2_{ge}}$) between MAGEPRO and LASSO across 1,000 independently simulated genes, where confidence intervals represent $\pm$ 1 standard error. Simulations were run with 4 causal variants shared across populations and true gene expression heritability of 10%. A black asterisk (*) above a pair of points indicates a significant difference of means ($p < 0.05$ from a two-sided unpaired t-test).

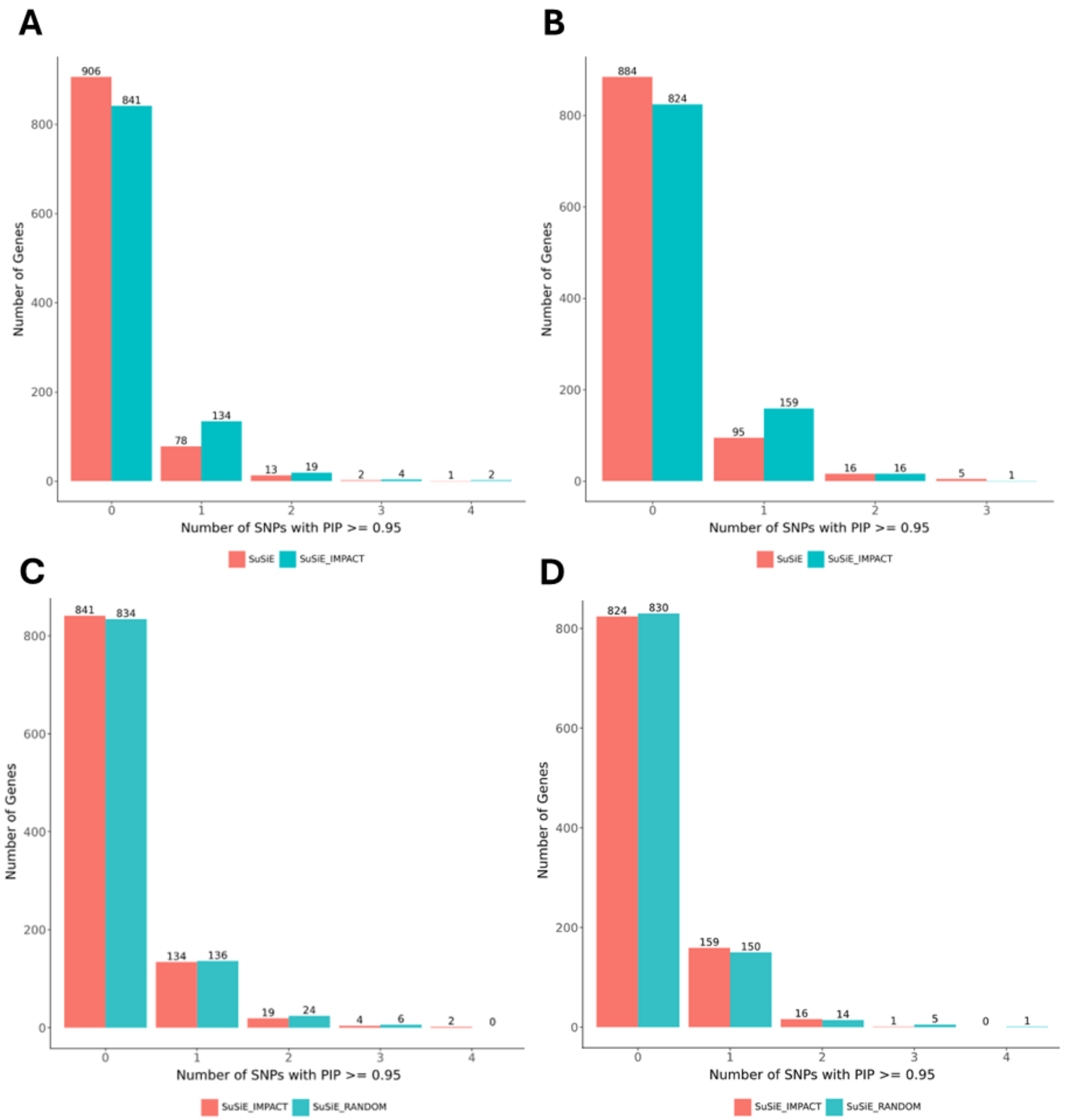


**Supplementary Figure 6. SNP-selection priors in SuSiE increases the number of genes with at least 1 putatively causal eQTL (PIP $\geq$ 0.95).**

(A) SuSiE with IMPACT functional annotations as SNP-selection priors applied to LCL gene expression from the GENOA AA population. (B) SuSiE with IMPACT functional annotations as SNP-selection priors applied to Monocyte gene expression from the MESA HIS population. (C) SuSiE with random SNP-selection priors applied to LCL gene expression from the GENOA AA population. (D) SuSiE with random SNP-selection priors applied to Monocyte gene expression from the MESA HIS population. PIP: posterior inclusion probability, LCL: lymphoblastoid cell line, AA: African American, HIS: Hispanic/Latino.

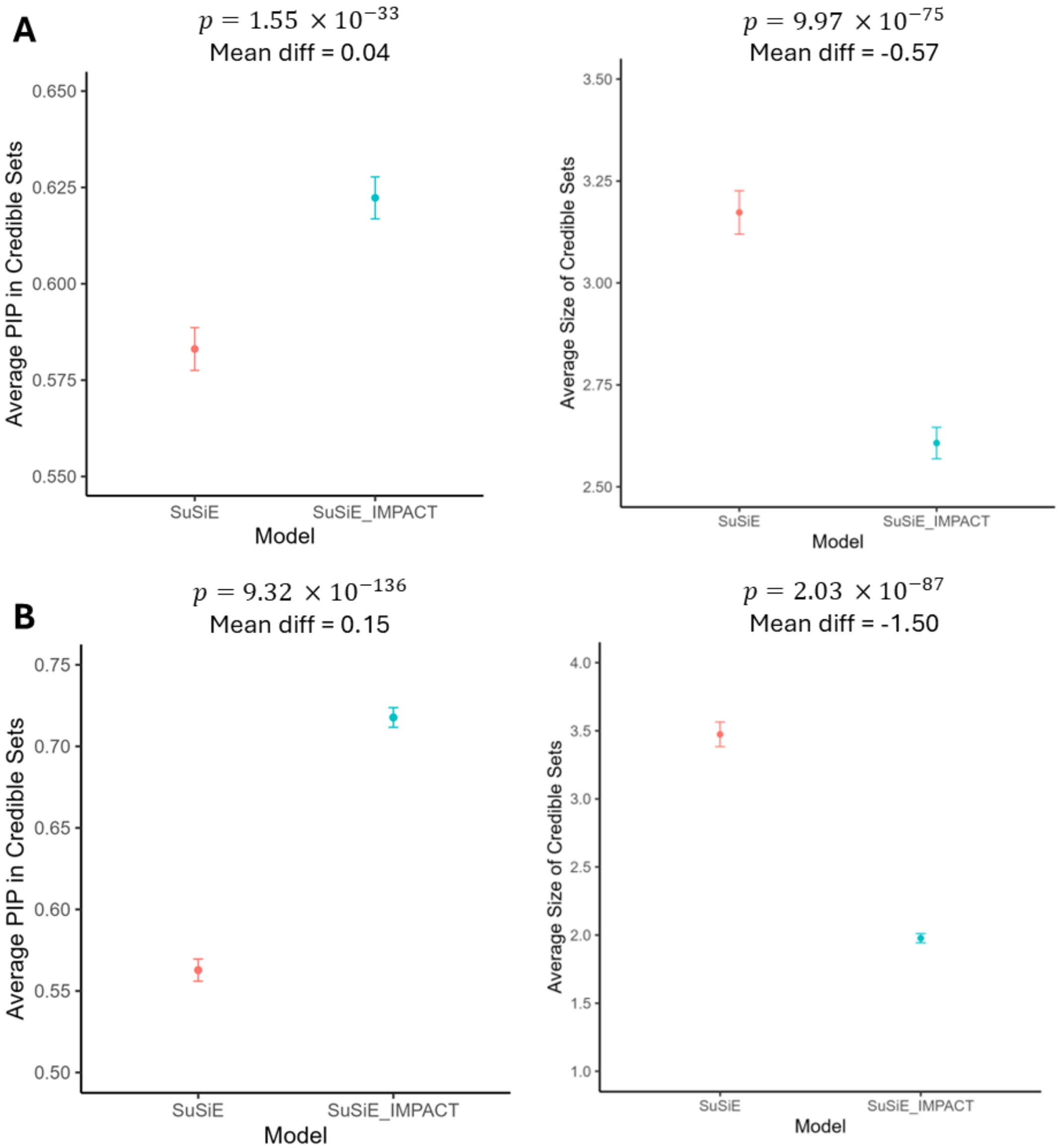


**Supplementary Figure 7. Leveraging IMPACT functional annotations as SNP-selection priors in SuSiE decreases the average credible set size and increases the average PIPs in credible sets.**

(A) Fine-mapping results from LCL eQTL summary statistics from GENOA AA. Average PIP of variants in 95% credible sets across the two approaches (left). Average number of variants in the 95% credible set across the two approaches (right). (B) Fine-mapping results from Monocyte eQTL summary statistics from MESA HIS. Average PIP of variants in 95% credible sets across the two approaches (left). Average number of variants in the 95% credible set across the two approaches (right). All hypothesis tests are two-sided paired t-tests and mean differences are relative to SuSiE without functional priors (SuSiE_IMPACT - SuSiE). In all panels, confidence intervals represent $\pm$ 1 standard error. PIP: posterior inclusion probability, LCL: lymphoblastoid cell line, AA: African American, HIS: Hispanic/Latino.

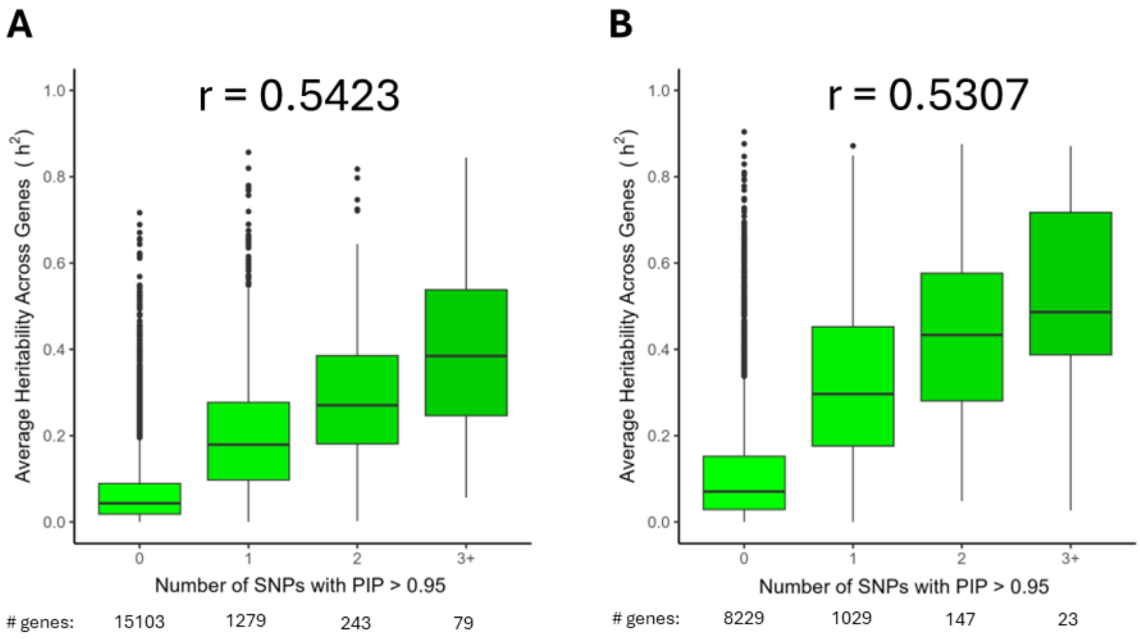


**Supplementary Figure 8. Genes with higher *cis*-heritability tend to have more putatively causal eQTLs.**

(A) LCL gene expression heritability from the GENOA AA population, estimated by GCTA. (B) Monocyte gene expression heritability from the MESA HIS population. In both panels, genes are partitioned by the number of putatively causal eQTLs identified by SuSiE (PIP $\geq$ 0.95). In both panels, data are represented by standard box and whisker plots. PIP: posterior inclusion probability, LCL: lymphoblastoid cell line, AA: African American, HIS: Hispanic/Latino.

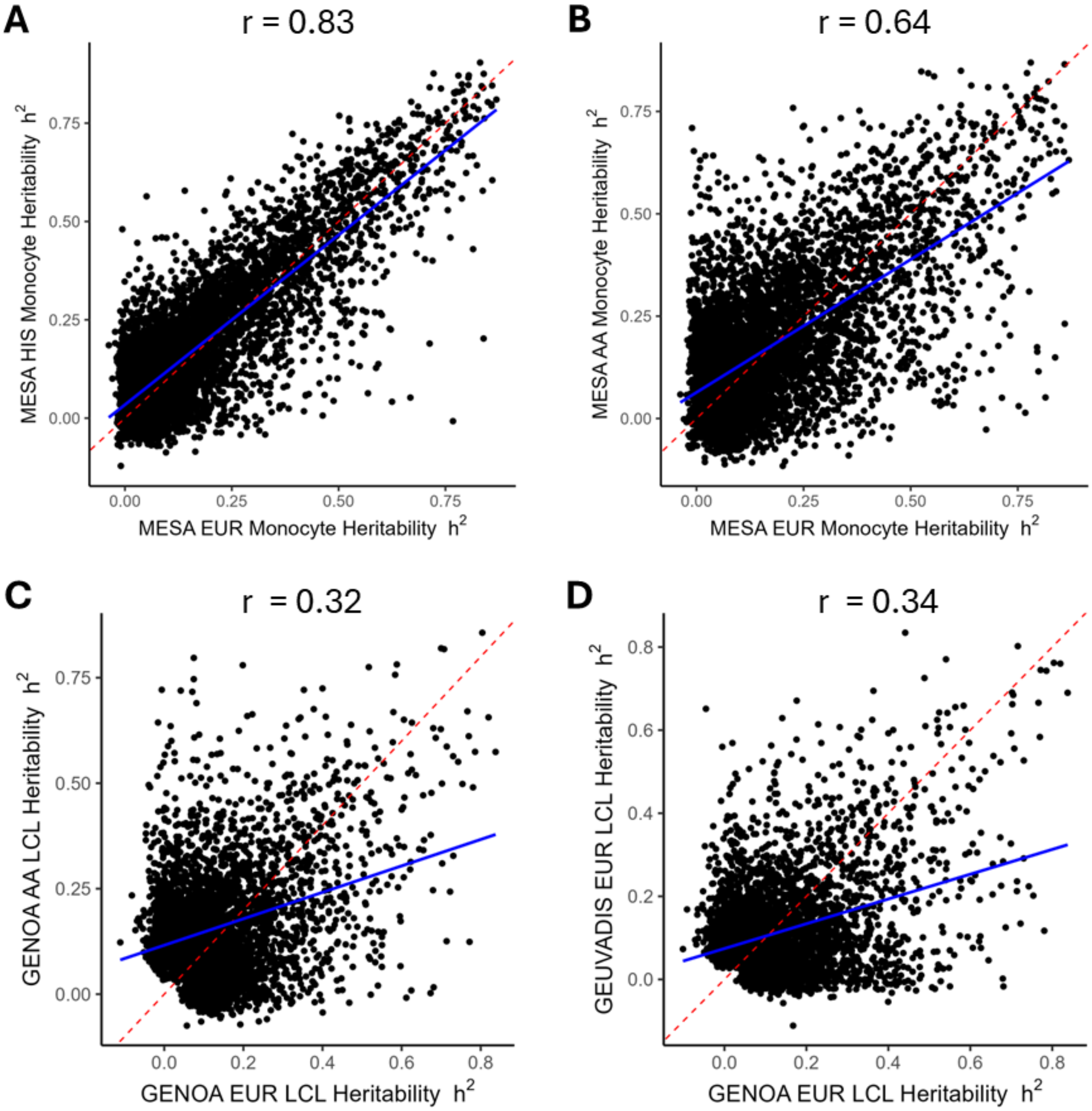


**Supplementary Figure 9. Gene expression *cis*-heritability is correlated across ancestries, but substantial cross-cohort heterogeneity exists within a given ancestry.**
(A) Comparison of gene expression heritability in monocytes between HIS and EUR individuals in MESA. (B) Comparison of gene expression heritability in monocytes between AA and EUR individuals in MESA. (C) Comparison of gene expression heritability in LCLs between AA and EUR individuals in GENOA. (D) Comparison of gene expression heritability in LCLs in EUR individuals between GEUVADIS and GENOA cohorts. In all panels, heritability estimates were computed with GCTA using only *cis*-SNPs that are shared between the two cohorts. Red dotted lines indicate $y = x$. Blue line indicates the line of best fit from linear regression. HIS: Hispanic/Latino; EUR: European; AA: African American; LCL: lymphoblastoid cell line.

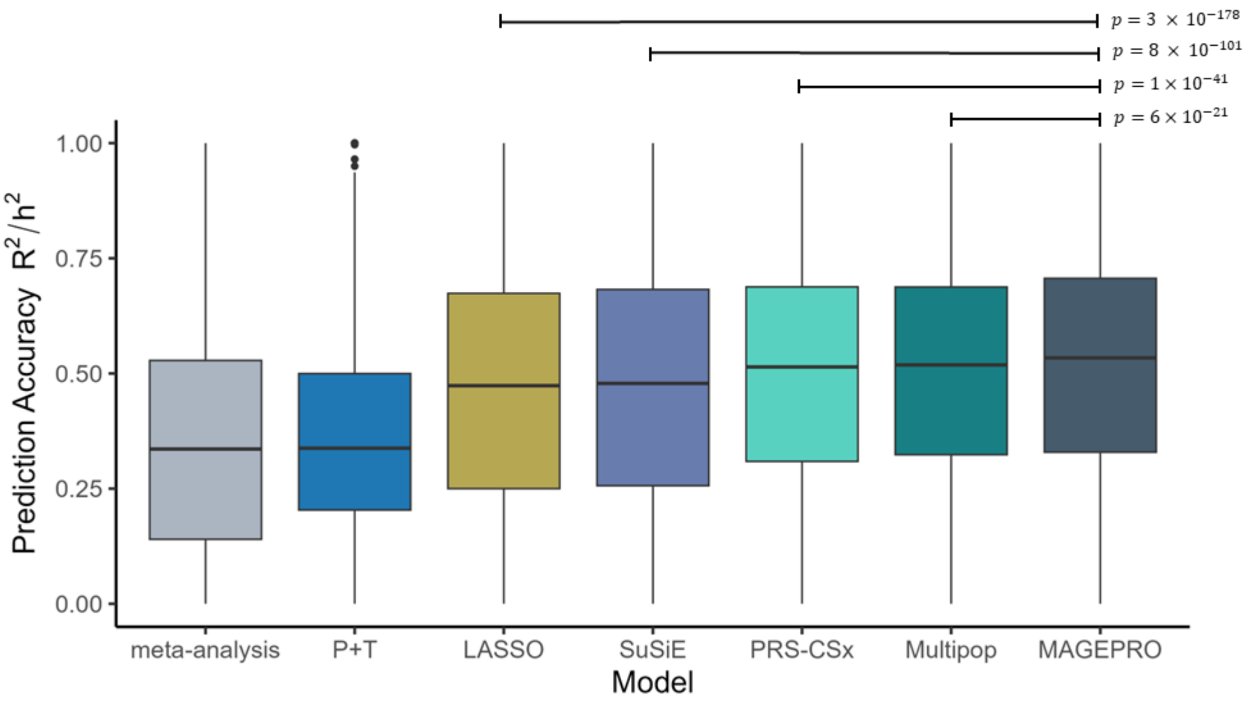


**Supplementary Figure 10. MAGEPRO outperforms alternative methods in predicting Monocyte gene expression in Hispanic/Latino individuals.**

Comparing accuracy ($\frac{R_{CV}^2}{\hat{h}_{ge}^2}$) of methods to predict Monocyte gene expression in the MESA HIS cohort. Data are represented by standard box and whisker plots. P-values are derived from paired one-sided t-tests. Comparisons between MAGEPRO and meta-analysis or P+T not annotated due to low precision to estimate such small p-values. HIS: Hispanic/Latino.

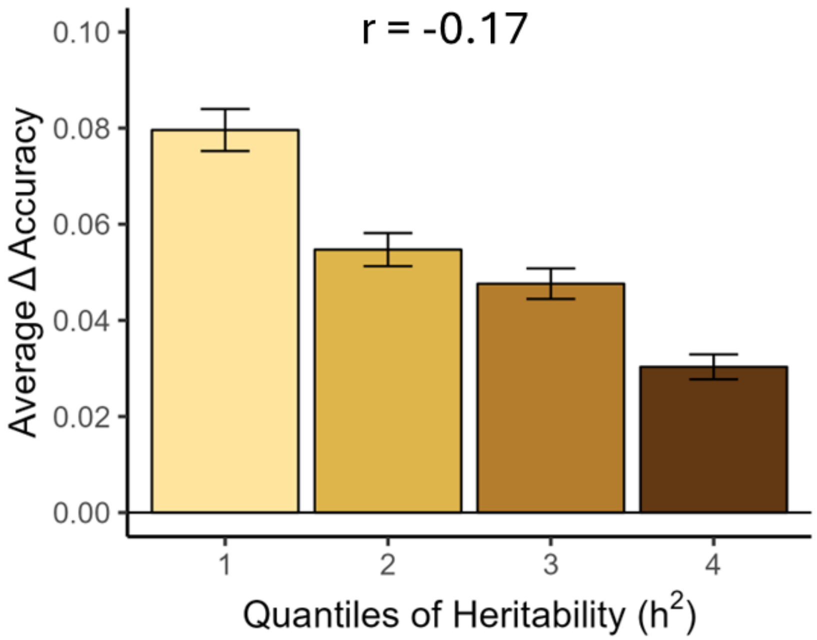


**Supplementary Figure 11. Change in accuracy between MAGEPRO and LASSO is negatively correlated with heritability estimates in MESA HIS cohort.**
Data plotted represent average change in accuracy (MAGEPRO $\frac{R_{CV}^2}{\hat{h}_{ge}^2}$ – LASSO $\frac{R_{CV}^2}{\hat{h}_{ge}^2}$) $\pm$ 1 standard error across 3,330 significantly *cis*-heritable genes. We note that while we show four quantiles here and 10 quantiles in **Figure 3D**, the correlation (r = -0.17) is computed over per-gene changes in accuracy and *cis*-heritability estimates. HIS: Hispanic/Latino.

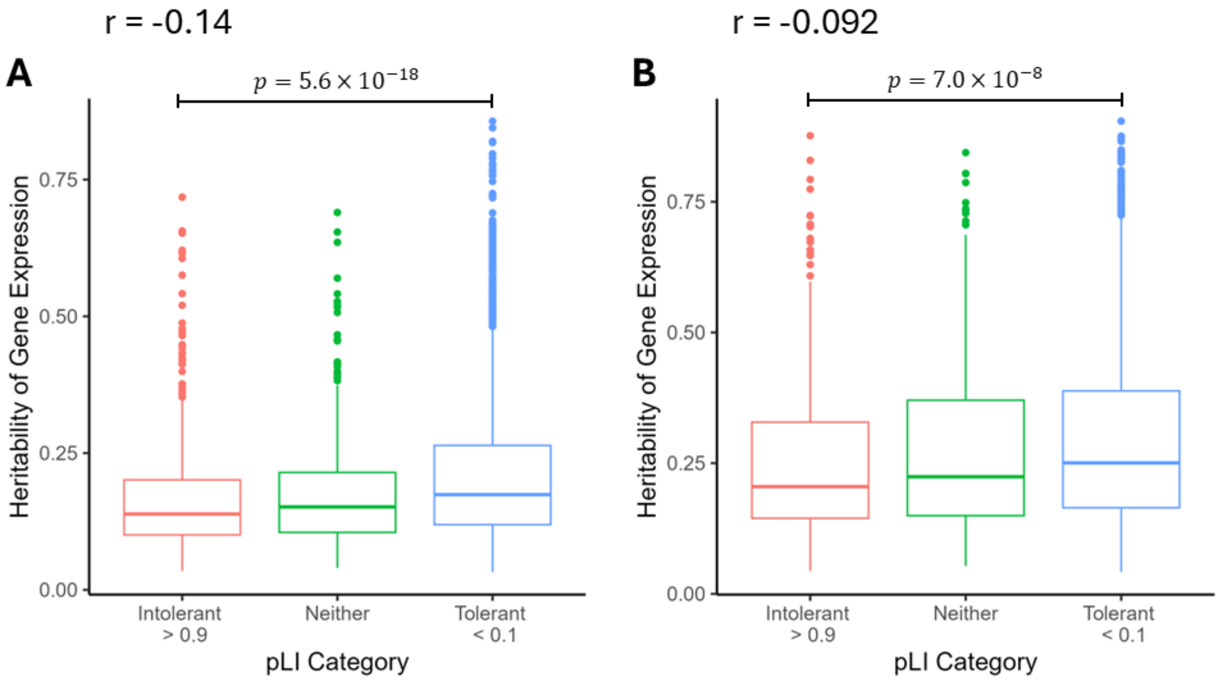


**Supplementary Figure 12. Loss-of-function intolerant genes have low heritability.** (A) Comparing the heritability of LCL gene expression in the GENOA AA cohort with gene pLI category. (B) Comparing the heritability of Monocyte gene expression in the MESA HIS cohort with gene pLI category. pLI cutoffs are based on previous literature (Lek et al. 2016 *Nature*). For both panels, Pearson correlation of heritability and pLI is computed across all significantly *cis*-heritable genes and data are represented by standard box and whisker plots. For the "neither" category, $0.1 \geq$ pLI $\leq$ 0.9. LCL: lymphoblastoid cell line; AA: African American; pLI: probability loss of function intolerant; HIS: Hispanic/Latino.

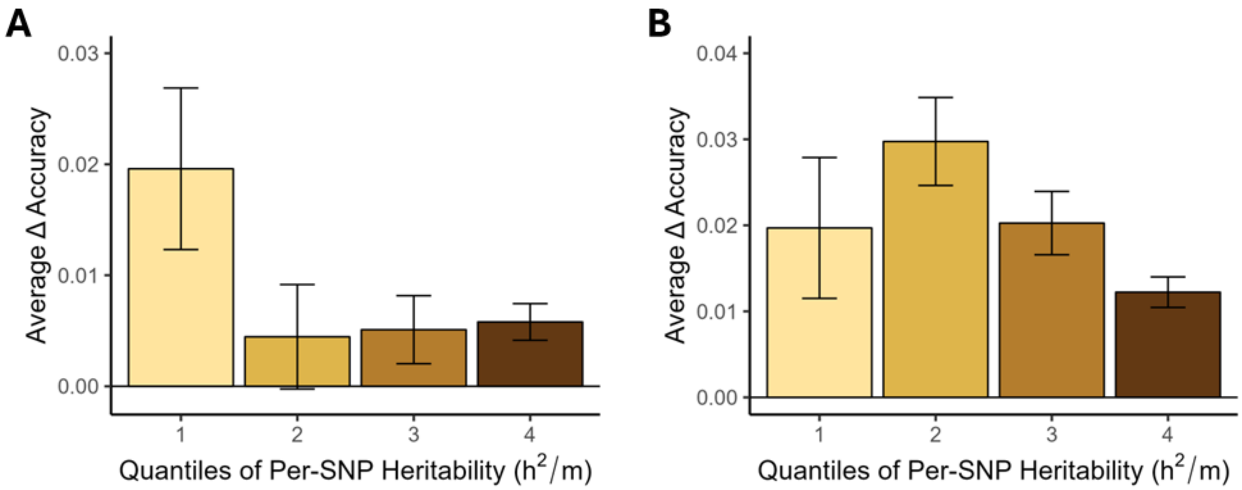


**Supplementary Figure 13. Change in accuracy between MAGEPRO and PRS-CSx is larger for genes with low per-SNP heritability.**

(A) Analysis in LCL gene expression data from the GENOA AA cohort; (MAGEPRO $\frac{R^2_{CV}}{\hat{h}^2_{ge}}$ – PRS-CSx $\frac{R^2_{CV}}{\hat{h}^2_{ge}}$) across four quantiles of gene expression *cis*-heritability. (B) Analysis in Monocyte gene expression data from the MESA HIS cohort. Per-SNP heritability was computed by normalizing the estimated heritability ($\hat{h}^2_{ge}$) by the number of putatively causal eQTL identified by SuSiE ($m = \#\, SNPs\ with\ PIP \geq 0.95$). Error bars represent average change in accuracy $\pm$ 1 standard error across significantly *cis*-heritable genes. LCL: lymphoblastoid cell line; AA: African American; HIS: Hispanic/Latino.

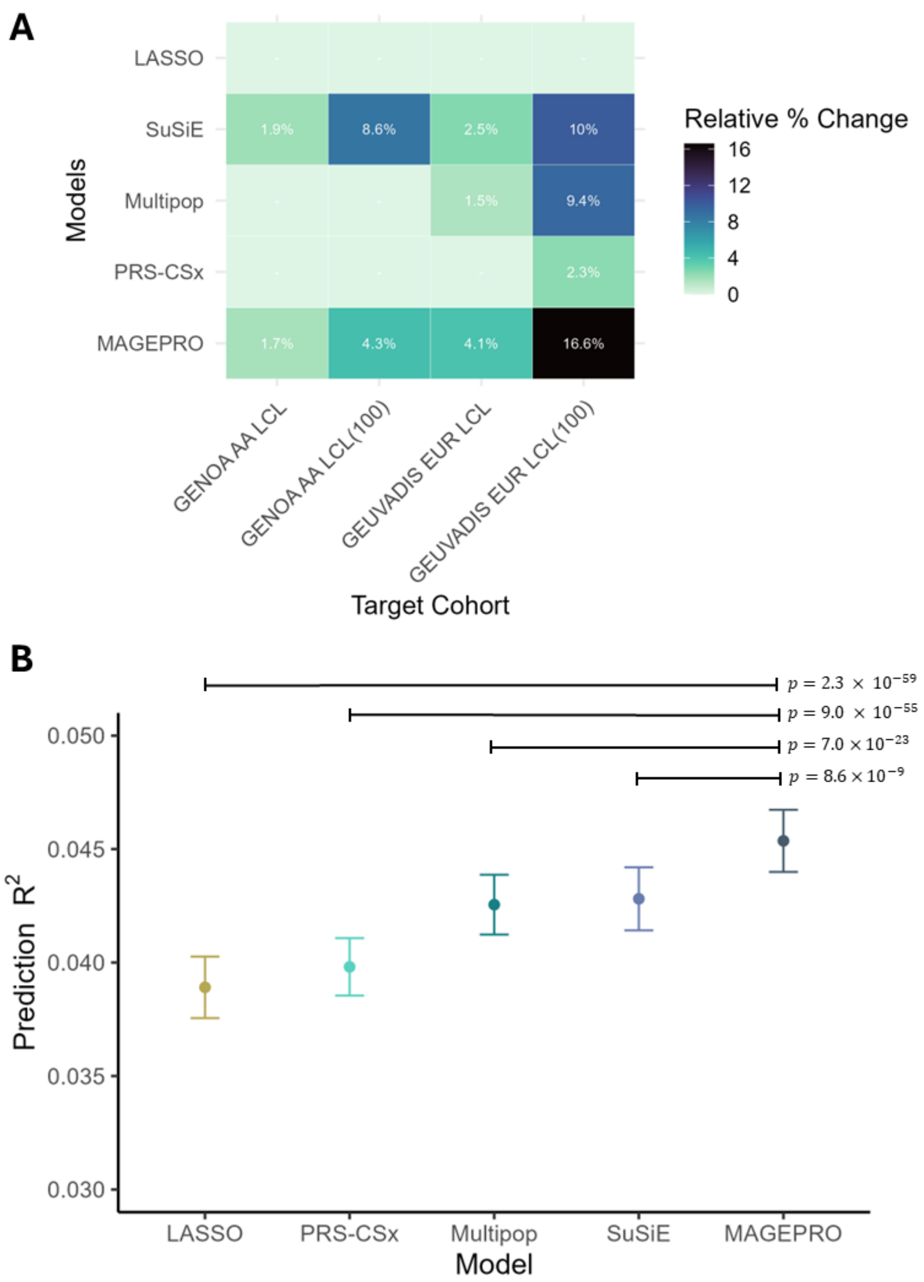


**Supplementary Figure 14. The relative performance of MAGEPRO in out-of-cohort prediction is variable.**

(A) Percent change in gene expression prediction $R^2$ relative to LASSO. Empty boxes indicate that there was no significant difference in prediction ($p > 0.05$ in a paired one-sided t-test). GENOA AA LCL models and GEUVADIS EUR LCL models were used to predict gene expression in the GEUVADIS YRI cohort and GENOA EUR cohort, respectively. (B) $R^2$ results from training models in downsampled GEUVADIS EUR ($n = 100$) and applying them to predict LCL gene expression in GENOA EUR cohort. Error bars represent average prediction $R^2$ $\pm$ 1 standard error across genes that were *cis*-heritable in either of the two cohorts ($\hat{h}^2_{in-sample} >$

0; $p < 0.05$ or $\hat{h}^2_{out-of-sample} > 0$; $p < 0.05$). Note, $R^2$ values are not directly comparable to previous figures reporting accuracy ($\frac{R^2_{CV}}{\hat{h}^2_{ge}}$). P-values are from a paired one-sided t-test. AA: African American; LCL: lymphoblastoid cell line; EUR: European; YRI: Yoruba.

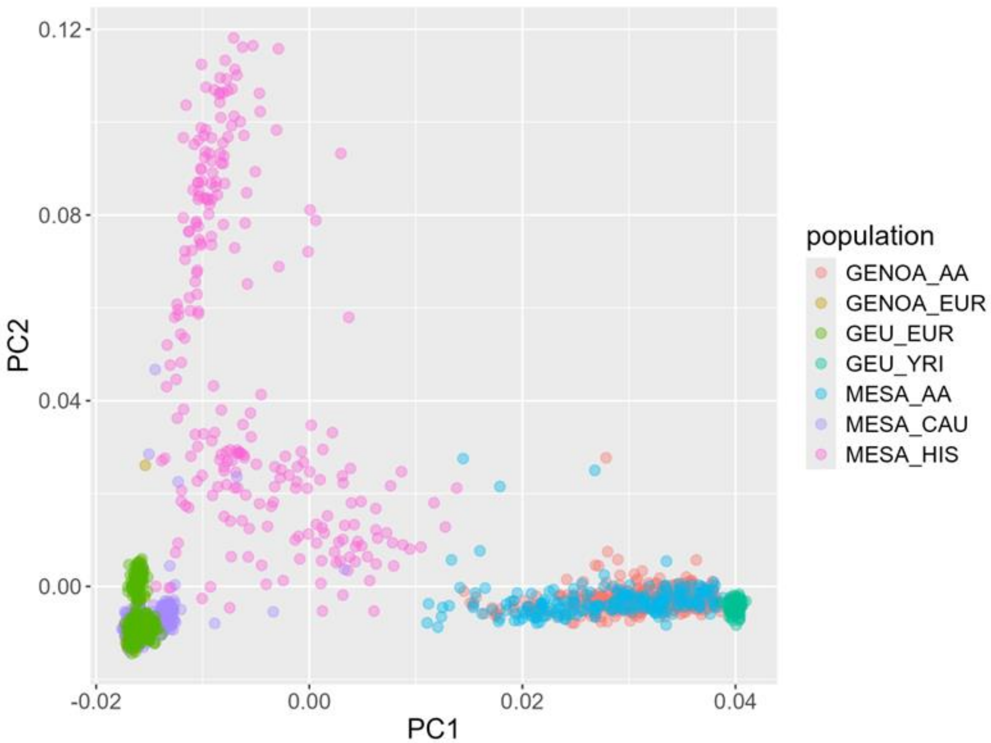


**Supplementary Figure 15. Genetic diversity of cohorts in this study.**
Principal components analysis was run on standardized genotypes for SNPs that were present in each of 7 distinct cohorts/populations discussed in the main text using PLINK. Here we visualize inter-individual variation across the first two principal components (PCs).

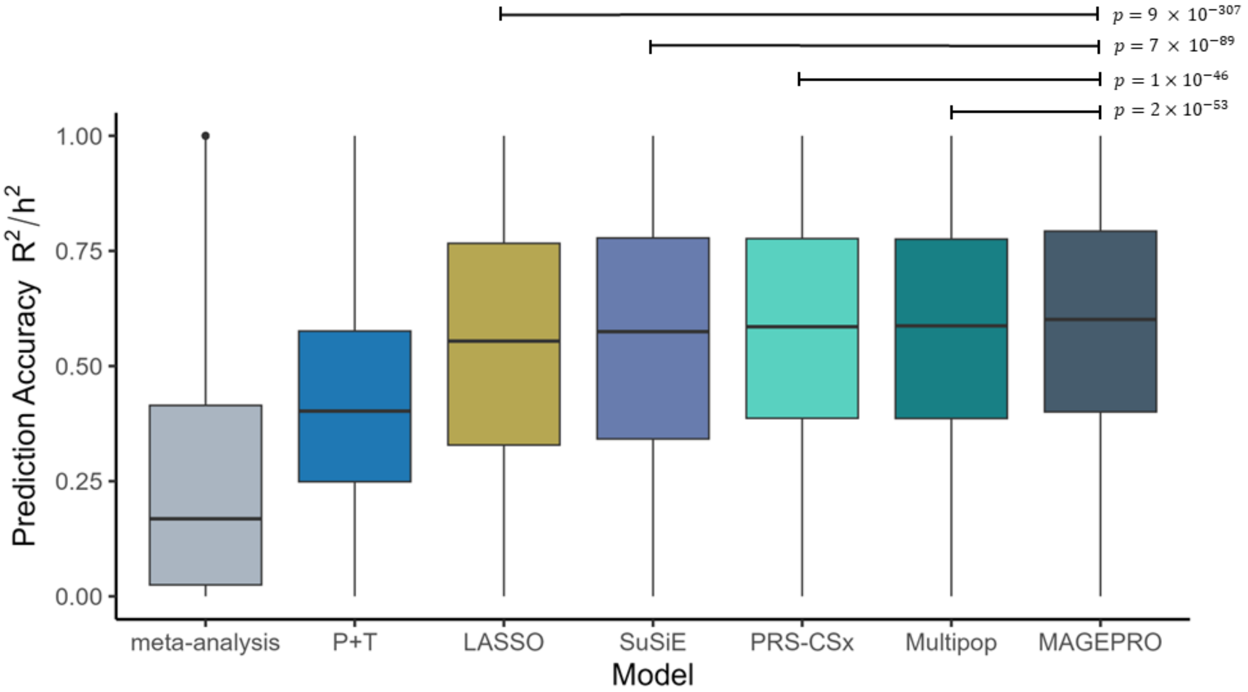


**Supplementary Figure 16. MAGEPRO outperforms alternative methods in predicting Lung gene expression in European individuals.**

Comparing the accuracy ($\frac{R^2_{CV}}{\hat{h}^2_{ge}}$) of methods to predict Lung gene expression in the GTEx EUR cohort. P-values are derived from paired one-sided t-tests. Data are represented by standard box and whisker plots. EUR: European.

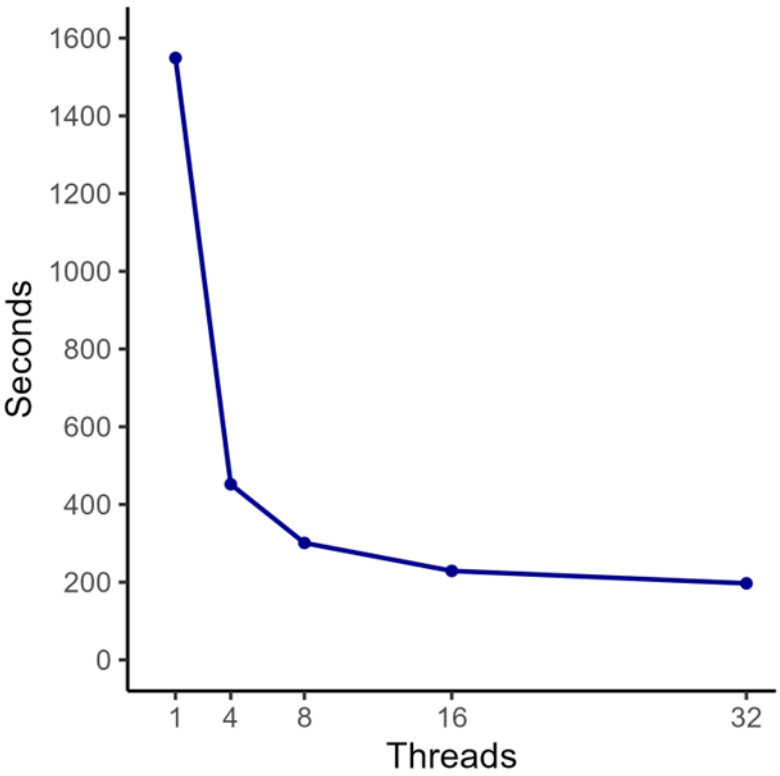


**Supplementary Figure 17. The runtime of MAGEPRO exponentially decays as the number of threads are increased.**

The runtime in seconds indicates the time it takes for our pipeline to (1) estimate gene expression heritability, (2) test LASSO and MAGEPRO in 5-fold cross validation, and (3) create genetic models of gene expression for 100 randomly selected genes.

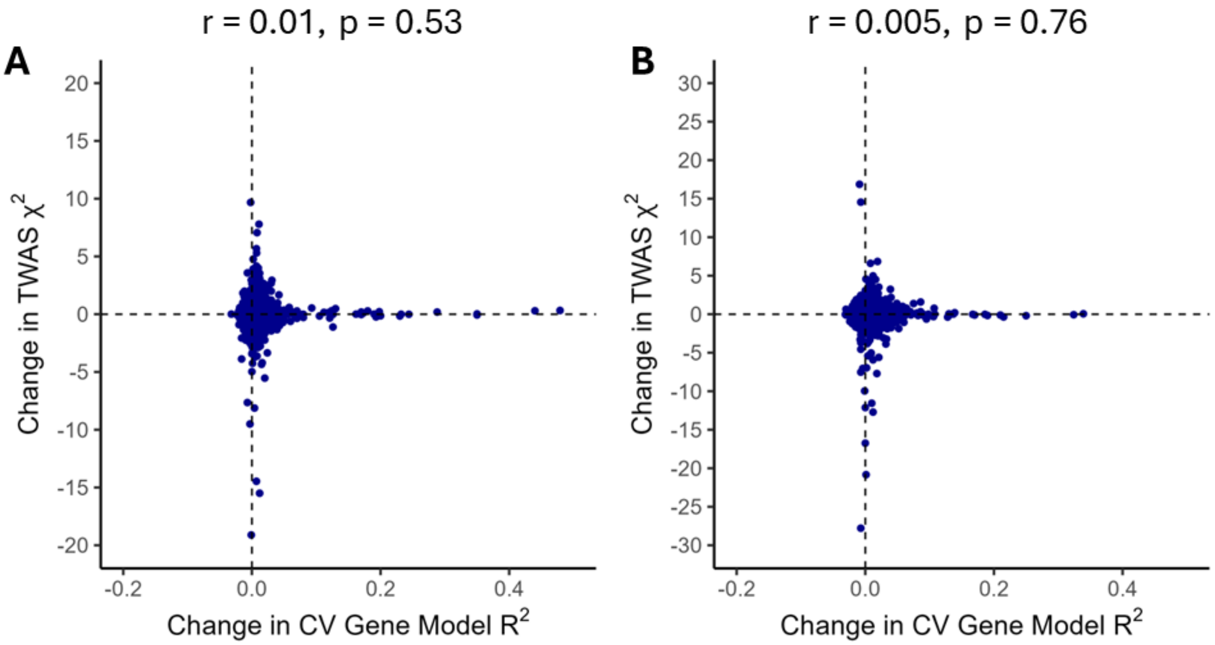


**Supplementary Figure 18. Improving gene expression prediction $R^2$ does not always strengthen TWAS associations.**

Relationship between change in gene expression prediction $R^2$ and change in TWAS $\chi^2$ (MAGEPRO - LASSO) for (A) LCL gene models trained in GEUVADIS EUR cohort applied to TWAS for Gout and (B) LCL gene models trained in GENOA AA cohort applied to TWAS for WBC count. CV: cross-validation; LCL: lymphoblastoid cell line; EUR: European; AA: African American; WBC: white blood cell.

| Cohort | Best | MAGEPRO | PRS-CSx | SuSiE | LASSO |
| --- | --- | --- | --- | --- | --- |
| MESA AA | 92 | 95 | 85 | 85 | 96 |
| MESA EUR | 6566 | 6608 | 6389 | 6564 | 6621 |
| MESA HIS | 7 | 6 | 7 | 6 | 5 |
| GENOA AA | 72 | 60 | 69 | 68 | 64 |
| GEUVADIS EUR | 4691 | 4670 | 4496 | 4691 | 4651 |
| GTEx AA | 33 | 27 | 39 | 26 | 29 |
| GTEx EUR | 8415 | 7225 | 8161 | 7131 | 6720 |

**Supplementary Figure 19. Number of significant TWAS associations found by each gene expression prediction method.**

Table displaying the number of significant TWAS associations found by each *cis*-genetic model of gene expression, aggregated across 22 unique traits/diseases. Gene models trained in each cohort were applied to ancestry-matched GWAS summary statistics. Genes with an $R^2$ significantly greater than 0 across all models are evaluated. “Best” refers to applying the gene model with the largest $R^2$ to TWAS. AA: African American; EUR: European; HIS: Hispanic/Latino.

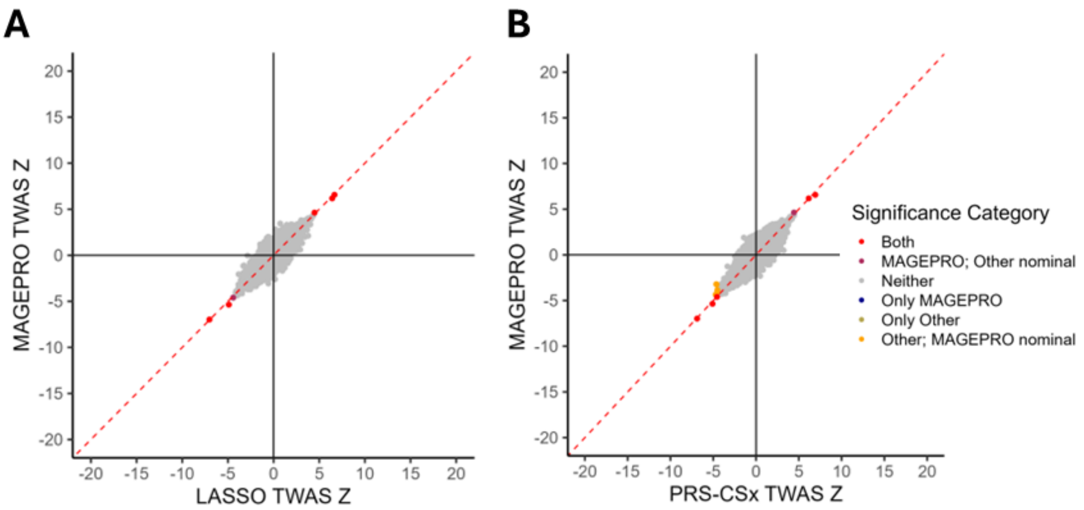


**Supplementary Figure 20. Comparison of gene expression prediction methods in TWAS for blood traits in a Hispanic/Latino population.**

Genetic models of gene expression were trained using Monocyte gene expression data from the MESA HIS (Hispanic/Latino) cohort for comparisons of (A) MAGEPRO to LASSO and (B) MAGEPRO to PRS-CSx. “Other” refers to the model in comparison on the x-axis. Results are aggregated across 15 blood cell traits.

#### ENSG00000116586 (*LAMTOR2*)

| Anc | %Acc LASSO | TWAS Z LASSO | %Acc MAGEPRO | TWAS Z MAGEPRO |
|---|---|---|---|---|
| EUR | 48% | **-7.3*** | 55% | **-7.1*** |
| AA | 28% | -3.1 | 44% | **-5.2*** |
| HIS | 31% | -3.4 | 39% | -3.7 |

#### ENSG00000116793 (*PHTF1*)

| Anc | %Acc LASSO | TWAS Z LASSO | %Acc MAGEPRO | TWAS Z MAGEPRO |
|---|---|---|---|---|
| EUR | 100% | **6.2*** | 100% | **5.92*** |
| AA | 0% ($R^2$ $not > 0$) | NA | 9% | **6.09*** |
| HIS | 14% | -0.89 | 20% | -0.56 |

#### ENSG00000134242 (*PTPN22*)

| Anc | %Acc LASSO | TWAS Z LASSO | %Acc MAGEPRO | TWAS Z MAGEPRO |
|---|---|---|---|---|
| EUR | 88% | **-6.88*** | 91% | **-7.07*** |
| AA | 59% | **-5.93*** | 59% | **-5.57*** |
| HIS | 45% | 1.45 | 45% | 1.47 |

#### ENSG00000160789 (*LMNA*)

| Anc | %Acc LASSO | TWAS Z LASSO | %Acc MAGEPRO | TWAS Z MAGEPRO |
|---|---|---|---|---|
| EUR | 74% | **-7.58*** | 74% | **-7.44*** |
| AA | 47% | **-5.75*** | 49% | **-4.79*** |
| HIS | 55% | -2.07 | 55% | -2.24 |

**Supplementary Figure 21. Examples of newly identified genes linked to white blood cell count, supported by recapitulated associations across multiple independent populations.** TWAS z-scores that are marked with asterisks are significant with Bonferroni correction ($p < \frac{0.05}{5920}$). TWAS z-scores labeled NA indicate that the gene was not tested due to an insufficient genetic model of gene expression ($R^2$ not significantly > 0). Gene expression prediction accuracy is calculated as $\frac{R^2_{CV}}{\hat{h}^2_{ge}}$. An accuracy of 100% may indicate that the gene model $R^2$ was greater than the *cis*-heritability. EUR: European; AA: African American; HIS: Hispanic/Latino.

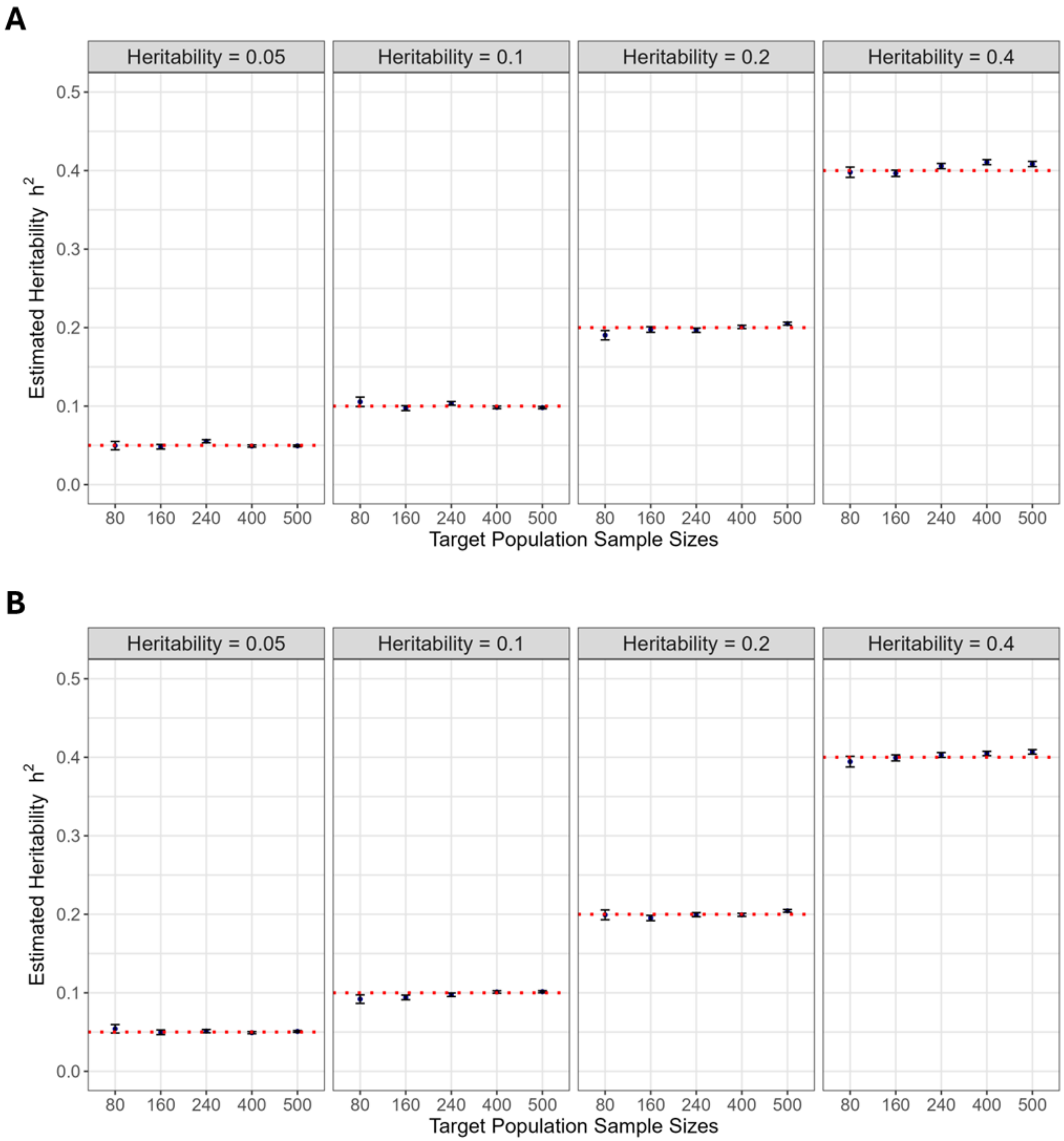


**Supplementary Figure 22. Simulation framework produces genes with desirable GCTA heritability estimates.**

(A) Heritability estimates for simulated gene expression when there is 1 causal variant shared across populations. (B) Heritability estimates for simulated gene expression when there are 4 causal variants shared across populations with correlated effect sizes. For both panels, red lines indicate the preset heritability value, and the data displayed are mean estimated heritabilities ± 1 standard error.

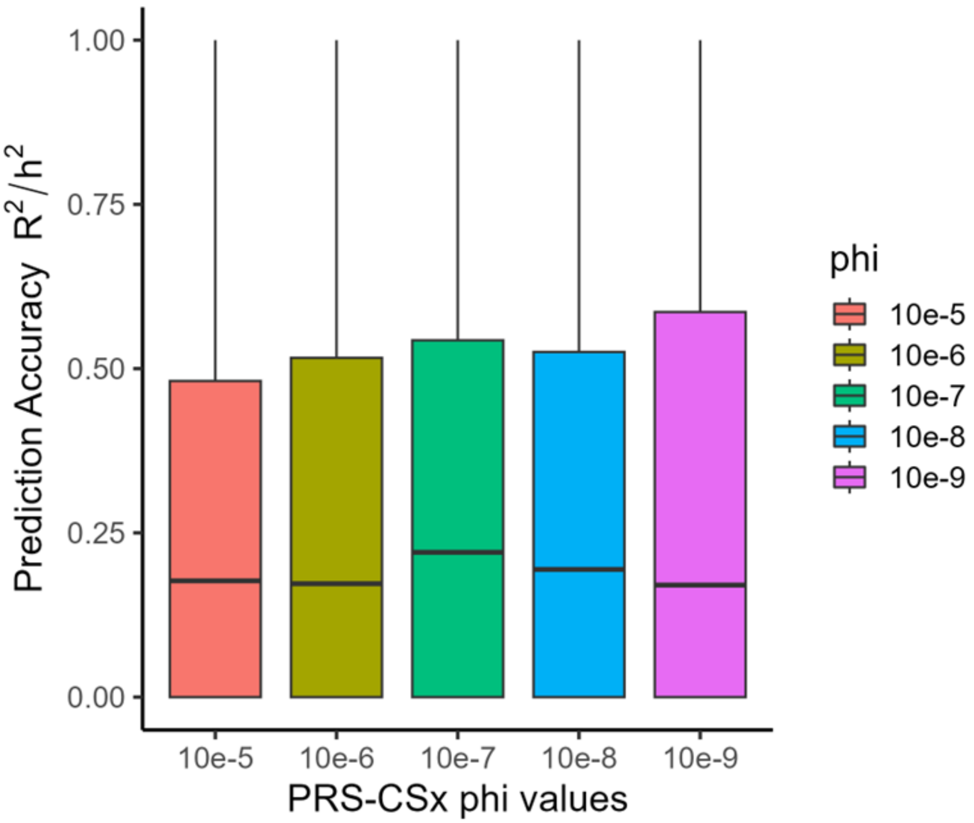


**Supplementary Figure 23. PRS-CSx model performance for different shrinkage parameters.**

We evaluated the accuracy ($\frac{R_{CV}^2}{\hat{h}_{ge}^2}$) of the PRS-CSx model across different shrinkage (phi, Φ) parameters for 200 randomly selected genes with $\hat{h}_{ge}^2 > 0$ and $\hat{h}_{ge}^2$ $p < 0.05$. While prediction accuracy was robust to any choice of phi (no statistically significant difference of means), we elected to set $\Phi = 10^{-7}$ as an intermediate value. We applied this value throughout the analysis of the remaining genes.

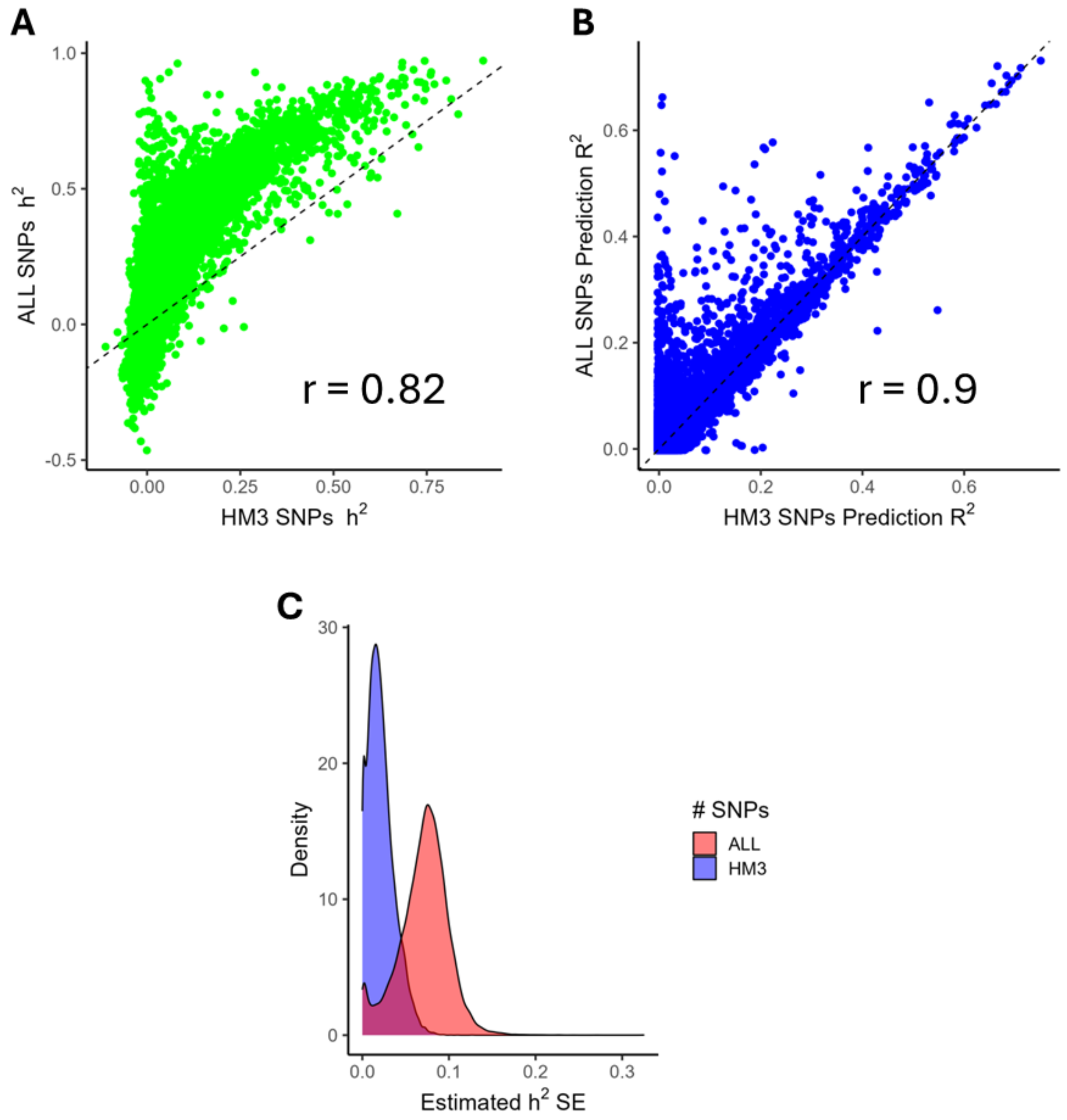


**Supplementary Figure 24. Comparison of heritability estimates and gene expression prediction $R^2$ when using all SNPs vs only HapMap 3 SNPs.**

(A) Comparison of estimated LCL gene expression heritability in the GEUVADIS EUR cohort when using only HM3 SNPs and when using all SNPs. (B) Comparison of LCL gene expression prediction $R^2_{CV}$ in the GEUVADIS EUR cohort when using only HM3 SNPs and when using all SNPs. (C) Distribution of standard errors of heritability estimates when using only HM3 SNPs and when using all SNPs. "ALL" SNPs refers to 38,187,570 SNPs present in the raw genotypes. EUR: European; LCL: lymphoblastoid cell line; HM3: HapMap 3.

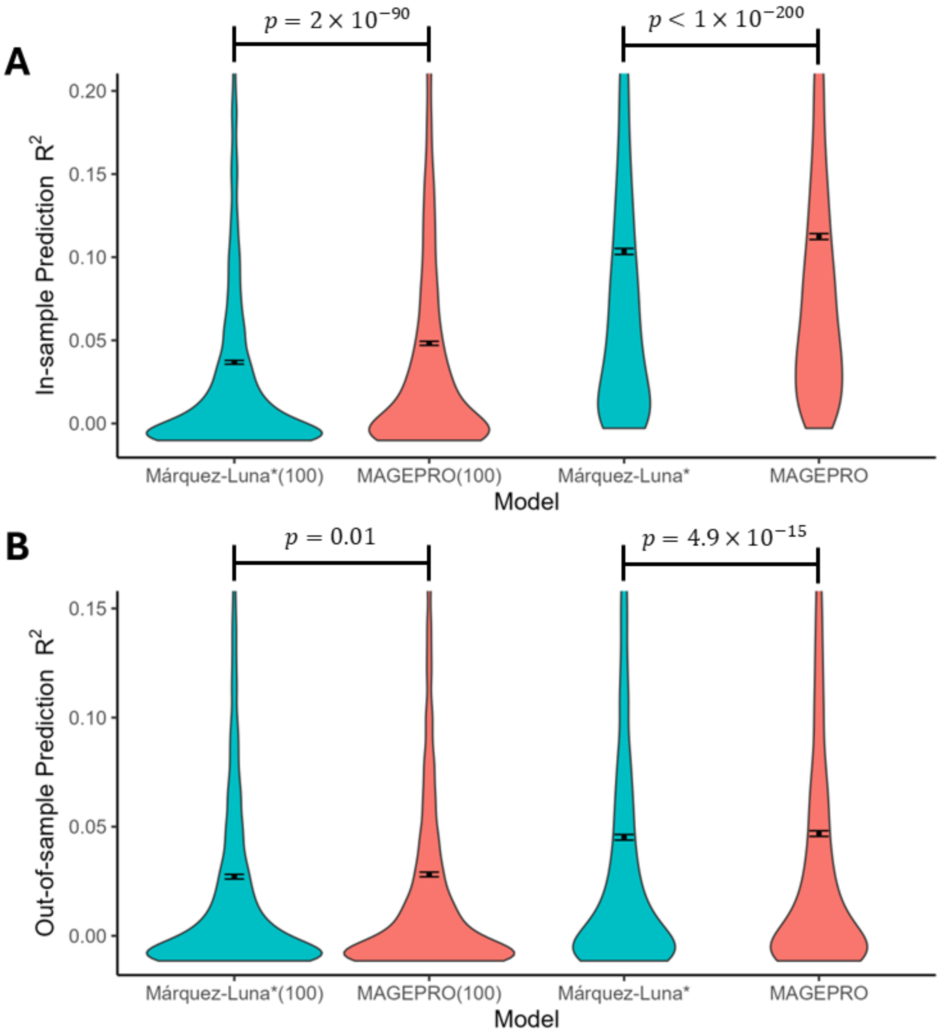


**Supplementary Figure 25. Comparison of in-sample prediction performance and out-of-cohort prediction performance for different training approaches.**
(A) In-sample prediction results of LCL gene models in the GENOA AA cohort from 5-fold cross validation. (B) Out-of-sample prediction results of LCL gene models trained in the GENOA AA cohort and tested in the GEUVADIS YRI cohort. All p-values are from a one-sided paired t-test (MAGEPRO > Márquez-Luna). “(100)” next to the model type indicates evaluation of the model when training samples are down-sampled to 100 individuals. AA: African American; LCL: lymphoblastoid cell line; YRI: Yoruba.
